# PhysioFusion: A Multi-Modal Ensemble using Static Preoperative Variables and Time Series Intraoperative Data to Predict Adverse Events Following Cardiothoracic Surgery

**DOI:** 10.64898/2026.09.24.26363920

**Authors:** Rajashekar Korutla, Anne Hicks, Marko Milosevic, Dipti Kulkarni, Felistas Mazhude, Tyler Kelting, Jaime B. Rabb, Qingchu Jin, Robert Kramer, Douglas Sawyer, Raimond L. Winslow, Saeed Amal

## Abstract

Accurate prediction of adverse events in the intensive care unit (ICU) following cardiothoracic surgery is crucial for timely interventions, improving patient outcomes and optimizing value in healthcare. By leveraging advanced machine learning techniques, this study demonstrates the transformative potential of predictive analytics to enhance postoperative care using a dataset from the Society of Thoracic Surgeons’ (STS) database [4] and time-series intraoperative data. We developed a multi-modal late fusion approach integrating static patient variables with intraoperative time-series data, utilizing an ensemble of models including residual neural networks for static features, CNN and Bidirectional GRU for time-series processing, XGBoost, logistic regression, SVM and Gradient Boosting. Through five-fold cross-validation, our ensemble model with balanced threshold selection achieved an AUC of 0.87, sensitivity of 0.76, specificity of 0.83, PPV of 0.40, and NPV of 0.96. Preoperative heart failure, IABP insertion, cardiopulmonary bypass (CPB) duration and white blood cell (WBC) count emerged as key predictors. Training the same pipeline on each modality separately showed that the static registry variables alone reached the same AUC of 0.87, and that the intraoperative signals alone reached 0.83, so on this cohort the two modalities act as substitutes rather than complements and fusion changed the sensitivity and precision trade-off rather than the amount of separable signal. A per-signal ablation found that no individual monitoring channel was load bearing, with mean arterial pressure and central venous pressure the least substitutable. This balanced approach optimizes the trade-off between sensitivity and specificity, providing a screening tool with high negative predictive value for monitoring high-risk patients, and the modality comparison indicates that comparable performance is attainable at sites holding registry data alone.

## 1 Introduction

### 1.1 Problem Definition and Motivation

Cardiothoracic surgery, despite advances in medical technology and practice, places patients at risk of morbidity and mortality. To prevent adverse events that lead to significant complications and death, timely and effective interventions are critical. To predict such events, a metric is necessary to analyze or predict their occurrence. Failure to rescue (FTR), defined as the mortality rate among patients who experience complications such as prolonged ventilation, stroke, renal failure or unplanned reoperation, is increasingly considered as a marker of hospital performance in caring for cardiac surgical patients post operatively [9, 22]. Studies [1] reveal significant disparities in risk-adjusted mortality rates following cardiac surgery, with differences as high as 2.5 times between low- and high-performing hospitals. To reaffirm this finding, hospitals with records from 2011 to 2018 were divided into three levels which showed an increase in FTR in a stepwise manner from low-mortality to high-mortality hospitals (8.3% vs 10.0% vs 12.7%, *P <* 0.001, respectively). This difference in FTR measures between hospitals with different rates of mortality underlines the opportunity for improved predictive models to alert the care team to potential complications before they occur [25]. Escalante et al. [2023] [23] utilized STS Adult Cardiac Surgery Database (ACSD) [4] data from 20,950 consecutive patients to demonstrate that the presence of cardiac-trained anesthesiologists, availability of extracorporeal membrane oxygenation (ECMO) and lower ratios of ICU beds to intensivists is associated with lower levels of FTR complications. This supports the assertion that matching appropriate resources to patient needs is essential to optimize patient outcomes and mitigate healthcare costs.

Stated concretely, the problem addressed in this study is the following binary prediction task. Given (i) the preoperative and procedural variables recorded for a patient in the STS ACSD and (ii) the physiological time series recorded during the operation, estimate, at the point at which the patient leaves the operating room, the probability that the patient will experience at least one of eleven STS National Quality Forum endorsed adverse events during the postoperative course. The eleven events are listed in Table 2 and are treated as a single composite outcome. Three properties of this task shape every methodological decision that follows. First, the outcome is rare: 161 of 1,393 patients in our cohort, or 11.6%, experienced at least one event, and the least frequent individual events occur in fewer than ten patients, which rules out reliable per-event modeling at this sample size. Second, the two available data sources are structurally different, one being a fixed-length vector of clinical and procedural descriptors and the other a set of variable-length physiological signals, so they cannot be concatenated naively. Third, the intended clinical use is screening rather than diagnosis: the model is meant to flag patients for closer observation at ICU handoff, so a high negative predictive value and a defensible operating point matter more than maximizing raw accuracy on an imbalanced dataset.

Three gaps motivate this work. The first is temporal: registry based risk models are built from preoperative information and are not designed to be updated once the operation has begun, even though key procedural exposures such as bypass and cross clamp duration are only known at its end. The second is informational: the intraoperative signals needed to close that gap are already recorded continuously for clinical monitoring, so using them adds no data collection burden and no additional cost or risk to the patient. The third is organizational: risk is currently carried across the operating room to ICU boundary by verbal handoff, during which information is known to be lost [40]. A model that consumes both data sources and emits a single calibrated probability addresses all three gaps at the point in the care pathway where the care team changes. The reported variation in failure to rescue rates across hospitals [9, 23] concerns mortality among patients who have already developed a complication, which is the stage of care that a pre-handoff risk estimate is intended to support.

### 1.2 Related Work

Prior work falls into five groups. For each we state what it does well and where it falls short for the task defined above, since these judgements motivated our design. Table 1 summarizes the positioning.

**Table 1:** Positioning of representative prior work with respect to the task defined in Section 1.1. Reported results are those stated by the cited studies; a dash indicates that no single comparable figure is reported in the source as cited here.

| Study | Data and cohort | Technique | Reported result | Strength | Limitation for the present task |
| --- | --- | --- | --- | --- | --- |
| Shahian, O’Brien [6, 5] | STS ACSD, national | Risk adjusted regression | Periodically recalibrated national models | National samples; inspectable form; accepted for consent discussions | Preoperative variables only; not designed for intra- or post-operative update |
| Ghaferi, Reddy [1, 3] | Medicare and cardiac surgery cohorts | Outcome epidemiology | Up to 2.5-fold mortality difference across hospitals | Establishes that outcomes differ where care differs, so events are partly recoverable | Hospital-level unit of analysis; no individual prediction |
| Kurlansky [9] | STS ACSD | FTR as a quality metric | Baseline for the present study | Gives the endpoint a registry endorsed definition | Quality measurement rather than pre-handoff alerting |
| Escalante [23] | STS ACSD, 20,950 patients | Association analysis | Staffing and ECMO availability associated with lower FTR | Identifies actionable structural factors at scale | Structural predictors; not patient-specific |
| Alabbadi [31] | NIS, 6,185,032 patients | Joinpoint regression | Significant FTR decline, 2000 to 2018 | Very large sample; quantifies secular trend | Administrative data, trend analysis, no intraoperative data |
| Bertsimas [21] | ECHSA, 295,000 operations | Optimal classification trees | Testing AUC 86.2% for mortality | Interpretable tree structure at very large scale | Congenital population; mortality endpoint; no physiological signals |
| Kilic [26] | STS National Database, 2007–2017 | XGBoost | Average observed-to-expected ratio 0.985 | Well calibrated against observed rates; handles non-linear interactions | Preoperative features; AVR population; separate models per outcome |
| Ebel [27] | In-hospital cardiopulmonary resuscitation | Artificial neural network | PPV 97% | Early evidence that neural networks can flag patients at risk of not surviving | Different setting and era; PPV not comparable across base rates |
| Ferrando-Vivas [28] | NHS cardiothoracic critical care, CMP | Refitted multivariable models | c-index 0.90 | Shows that adding intraoperative data raises discrimination | Summarized intraoperative variables; mortality endpoints |
| Fernandes [29] | Cardiac surgery, preoperative plus intraoperative | XGBoost | Hypotension outside CPB improved discrimination | Localizes the signal to a specific intraoperative exposure | Hand-derived exposure summaries; mortality endpoint |
| Penny-Dimri [20] | Meta-analysis, 51 studies | Bayesian pooling of C-indices | Pooled C-index 0.82 for ML vs 0.80 for LR, 30-day mortality | Supplies a field-level benchmark with credible intervals | Mortality endpoints only; no significant ML gain over LR |
| Sulague [32] | Systematic review, 81 studies | Survey of AI methods in cardiac surgery | Random forest, SVM, LR and XGBoost most common | Maps which methods and procedures the field has covered | Reports coverage rather than performance; safety not yet established |
| Guo, Revathi, Hossain, Ogunpola [16, 14, 18, 17] | EHR and clinical time series, non-surgical | LSTM, CNN-LSTM with explainability, comparative ML | — | Architectures suited to local morphology and long dependencies | Non-surgical cohorts; single modality; larger samples |
| Bhattacharya [2] | Post-PCI cardiovascular outcomes | Multi-modal fusion | — | Applies late fusion of modalities to an adjacent cardiovascular outcome | Different procedure and endpoint; no per-signal ablation |
| This work | STS ACSD paired with 26 intraoperative signals, 1,393 patients | Late fusion ensemble with per-signal ablation | AUC 0.87, sensitivity 0.76, specificity 0.83, NPV 0.96 | Uses raw signals, compares modalities, attributes value per channel, calibrated | Single center; moderate case count; composite endpoint |

#### Registry based risk models

The cardiac surgical community relies on the STS ACSD to estimate risk adjusted outcomes and to counsel patients preoperatively [6, 5]. These models are fitted on national samples, risk adjusted, and expressed in regression forms clinicians can inspect [7]. Their limitation for our task is structural: they consume only preoperative information and are not designed to update a risk profile intra- or post-operatively, so a patient whose operation went badly leaves the operating room carrying the same predicted risk as one whose operation went smoothly.

#### Epidemiological and quality metric studies of failure to rescue

A second group establishes that the outcome is worth predicting. Ghaferi et al. [1] reported risk-adjusted mortality differences as high as 2.5 times between low- and high-performing hospitals, Reddy et al. [3] framed FTR as an improvement opportunity in cardiac surgery, Kurlansky et al. [9] formalized it as an STS quality metric, Milojevic et al. [19] and Likosky et al. [24] documented interhospital variation, and Escalante et al. [23] showed in 20,950 patients that cardiac-trained anesthesiologists, ECMO availability and lower ICU bed to intensivist ratios are associated with lower FTR. Alabbadi et al. [31] applied joinpoint regression [37] to 6,185,032 National Inpatient Sample patients [36] and reported a decline in FTR from 2000 to 2018, and broader reviews of FTR [25] and of complication epidemiology in esophagectomy [30] reach similar conclusions. The strength of this literature is scale and credibility; its limitation is that the unit of analysis is the hospital or the era rather than the patient, and the designs are associational. It justifies building a predictive tool but does not supply one.

#### Machine learning on preoperative and administrative data

Kilic et al. [26] trained XGBoost on STS National Database [35] entries from 2007 to 2017 for operative mortality, renal failure and deep sternal wound infection, reporting an observed-to-expected ratio of 0.985. Bertsimas et al. [21] used 295,000 operations from the ECHSA Congenital Database [34] and found optimal classification trees best, with a testing AUC of 86.2% for mortality. Ebel [27] applied neural networks to failure to survive after in-hospital resuscitation, reporting a PPV of 97%. Tree based ensembles here capture non-linear interactions while exposing feature importance, and large samples give narrow intervals. Three limitations matter: the predictors remain preoperative or administrative, so the operation is a black box; the endpoints are mortality or one named complication rather than a composite of recoverable events; and headline metrics are not comparable across base rates, since a PPV of 97% in a high-prevalence cohort [27] does not transfer to one with an event rate of 11.6%. The field’s own syntheses temper these results. Penny-Dimri et al. [20] screened 2,792 references, included 51 studies, and pooled a C-index of 0.82 (95% credible interval 0.79 to 0.85) for machine learning against 0.80 (0.77 to 0.84) for logistic regression and 0.78 (0.74 to 0.82) for scoring tools on 30-day mortality, with 0.81 against 0.79 for in-hospital mortality, finding no statistically significant superiority for machine learning and noting that such models have not reached routine use. Sulague et al. [32] identified 81 studies with a rapid rise since 2020, dominated by random forests, support vector machines, logistic regression and extreme gradient boosting, applied mainly to preoperative risk assessment, and concluded that accuracy and safety need further verification; Salna [33] argues that the value of these methods spans the surgical journey and depends on interdisciplinary collaboration. The lesson is cautionary: gains from algorithmic sophistication alone appear small, so a new model is worth reporting only if it adds information the existing predictors do not carry, states performance in a comparable form, and is evaluated at an operating point corresponding to a real decision.

#### Studies that add intraoperative information

Ferrando-Vivas et al. [28] refitted models for acute hospital and 1-year mortality after cardiothoracic critical care admission, adding pre- and intra-operative data from NHS units in the Case Mix Programme, reaching a c-index of 0.9. Fernandes et al. [29] predicted postoperative mortality from mean arterial pressure, vasopressor and inotrope administration, surgical duration and cross-clamp time alongside preoperative risk factors, finding with XGBoost that intraoperative hypotension outside the bypass phase gave better discrimination, sensitivity, specificity and PPV than the comparison models. The premise of our paper follows from this group. Its limitations are equally instructive: both consume the intraoperative record as a few hand-derived summaries, which requires knowing in advance which aspect of each signal matters; both target mortality rather than the recoverable events that define FTR; and neither reports what each individual signal contributes, nor the preoperative-only comparison on the same folds that would isolate the value of the added data.

#### Deep learning on clinical time series, and multi-modal fusion

Recurrent models have been applied to longitudinal records, including LSTMs for cardiovascular health trajectories [16] and disease prediction [14], and hybrid convolutional and recurrent models with explainability components [18]. Boosted trees and comparative model studies address related clinical risk problems including hypertension outcomes [13], infection risk in cirrhosis [15] and cardiovascular disease detection [17]. On fusion, Bhattacharya et al. [2] applied a multi-modal fusion model to adverse cardiovascular outcomes after percutaneous coronary intervention, and work on multi-modal cardiovascular data [10], usability testing of deployed screening tools [11, 12] and explainable pathway visualization [38] addresses what happens after a model is built. Convolutional layers suit local morphology and recurrent layers carry longer dependencies, and the implementation studies show a model only changes care if clinicians can act on it. The limitations are that these architectures are demonstrated on cohorts far larger than a single-center surgical registry, that most are single-modality or non-surgical, and that calibration, which determines whether a predicted risk can be read as a risk [8], is reported less often than discrimination.

#### Lessons learned from the literature

Seven conclusions follow from the studies above. Each is stated as a lesson, followed by the design choice it produced in this study and the guidance we would offer a researcher approaching the same problem.

1. **The operation carries predictive information that preoperative models discard.** The two studies that incorporated intraoperative variables report strong discrimination with them, a c-index of 0.90 in [28] and, in [29], better discrimination, sensitivity, specificity and PPV for models using intraoperative hypotension outside the bypass phase than for the comparison models reported there. We therefore paired the STS variables with the signals recorded during the same operation. *Guidance:* before adding architectural complexity, check whether an unused data source is available, since a new modality has repeatedly produced larger gains than a new algorithm in this problem.
2. **Reducing signals to hand-picked summaries presupposes the answer.** Where the intraoperative record has been used, it was compressed into a few derived exposures such as durations and hypotension burden [28, 29], which requires knowing in advance which aspect of each channel matters. We therefore passed the signals to learned encoders, convolutional for local and transient behaviour and bidirectional recurrent for longer dependencies. *Guidance:* keep at least one configuration that learns from the raw signal, so that the summary choice can be tested rather than assumed.
3. **Model family should be chosen against sample size, not fashion.** Tree based models are the reported performers in the structured clinical data studies cited above [26, 21, 15], while the recurrent and hybrid architectures that excel on physiological sequences are demonstrated on cohorts far larger than a surgical registry [16, 14, 18]. With 161 cases we therefore combined tree based, linear, margin based and deep models rather than relying on one family. *Guidance:* at case counts in the low hundreds, treat a deep sequence model as one voice in an ensemble rather than the primary estimator, and report what it contributes separately.
4. **Algorithmic complexity has to be earned.** A meta-analysis of 51 studies found no statistically significant discrimination advantage for machine learning over logistic regression for mortality after cardiac surgery [20]. We therefore retained logistic regression as a component model rather than as a straw man and report the margin between it and the full ensemble for every configuration in Sections 4.1 to 4.3. *Guidance:* always publish a regularized regression baseline on the same folds; if the margin is small, that is itself the finding.
5. **The endpoint should match the clinical question.** Prior models predict mortality or one named complication [26, 21, 28, 29], which is not what failure to rescue concerns. We therefore predicted the composite of eleven STS National Quality Forum endorsed events. *Guidance:* a composite buys enough events to fit a model but costs interpretability, so state the trade-off explicitly and report the component counts, as in Table 2.
6. **Headline metrics are not portable across cohorts.** A positive predictive value of 97% [27] and an observed-to-expected ratio of 0.985 [26] cannot be read beside a PPV obtained at an 11.6% event rate, because PPV depends on prevalence. We therefore report sensitivity, specificity, PPV, NPV, F1 and AUC with confidence intervals at explicitly named thresholds. *Guidance:* report the event rate next to every prevalence-dependent metric, and prefer AUC or C-index when comparing across cohorts.
7. **Discrimination alone does not make a model usable.** Calibration determines whether a predicted risk can be read as a risk [8], whereas the pooled evidence for this problem is expressed in discrimination alone [20], and the field’s own review concludes that accuracy and safety require further verification before clinical use [32]. Implementation studies show that a model changes care only when clinicians can act on its output [11, 12, 38]. We therefore applied isotonic calibration, reported calibration error alongside discrimination, and selected the operating point for a screening use case. *Guidance:* fix the intended decision first, then choose the threshold and the metrics that decision requires, rather than reporting performance at a default cutoff.

**Table 2:** Counts of target variables in the dataset.

| Target Name | Positive Counts |
| --- | --- |
| Unplanned post operative aortic re-intervention | 2 |
| Unplanned post operative reoperation for valvular dysfunction | 4 |
| Post operative deep sternal wound infection | 5 |
| Unplanned post operative coronary artery intervention | 7 |
| Unplanned post operative reoperation for other cardiac reasons | 19 |
| Permanent post operative neurologic deficit | 25 |
| Post operative renal failure | 39 |
| Reoperation for bleeding / cardiac tamponade | 46 |
| Discharged from hospital deceased | 46 |
| Deceased within 30 days of surgical intervention | 49 |
| Prolonged post operative mechanical ventilation | 105 |
| **Total occurrences** (including patients with multiple occurrences) | **161** |

Taken together, the literature leaves three specific gaps that this study addresses: no prior study in this setting combines registry variables with the raw intraoperative signal set for the FTR-defining composite of events, none reports what each individual monitoring channel contributes to such a prediction, and few report calibrated probabilities at a clinically named operating point. It also supplies the yardstick against which our results should be read, namely a pooled C-index of about 0.82 for mortality prediction after cardiac surgery [20], with the caveat that our endpoint is a composite of eleven events rather than mortality and our cohort is a single center, so the comparison is indicative rather than like for like. These gaps define the contributions listed in the next section.

### 1.3 Proposed Solution and Contributions

We address the task with a multi-modal late fusion ensemble that keeps the two data sources separate for as long as their structures differ and combines them only at the level of learned representations and model outputs, providing a further mechanism to communicate risk to the ICU team. The pipeline, whose deep learning component is shown in Figure 1 and whose details are given in Section 2, proceeds in five stages. It consumes the static vector of STS ACSD variables together with 26 intraoperative signals covering arterial, aortic and central venous pressures, pulse rate, electrocardiographic leads, oxygenation, temperature, arterial flow and bypass timings. Each modality is preprocessed on its own terms, with type-specific transforms for the static variables and padding with robust scaling for the signals. Representations are then learned in parallel branches, a residual network for the static features and, over the signals, a convolutional pathway for local and transient behaviour alongside a bidirectional GRU pathway for longer dependencies, while XGBoost, L1-regularized logistic regression, a support vector machine and a two-stage gradient boosting cascade are trained on the same inputs so that tree based, linear and margin based decision boundaries are all represented. The branch representations are concatenated and the component predictions combined with optimized weights, with isotonic regression applied so that scores can be read as probabilities. Finally, because the intended use is screening, the operating point is chosen explicitly, and we evaluate PPV-optimal, F1-optimal and balanced thresholds under five-fold cross-validation with resampling, reporting calibration error alongside discrimination.

**Figure 1:**
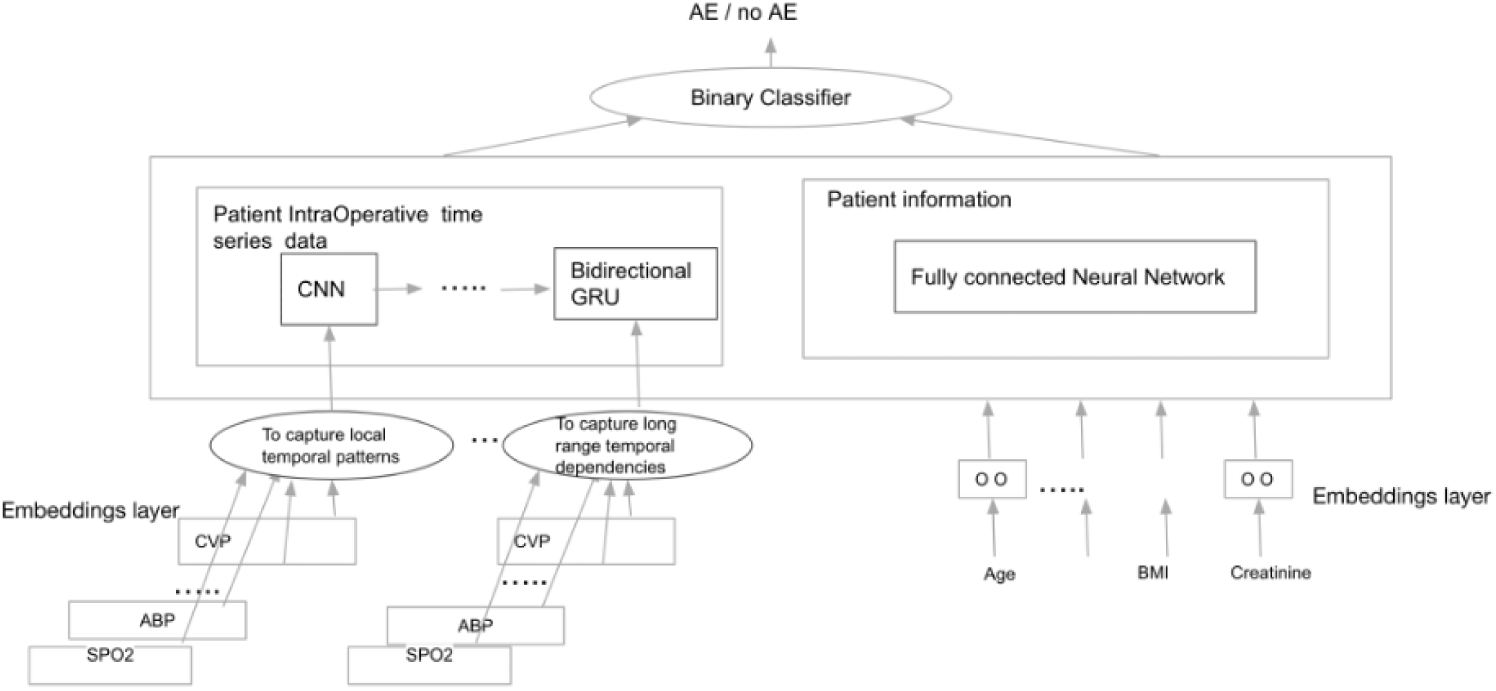
Deep learning architecture within the ensemble model.

To separate what the intraoperative record adds from what the registry already provides, this identical pipeline is trained on static features alone, on time series alone, and on both, and the combined configuration is then subjected to a per-signal ablation in which one time series variable at a time is withheld. With the balanced threshold, the combined ensemble reached an AUC of 0.87, sensitivity of 0.76, specificity of 0.83, PPV of 0.40 and NPV of 0.96, with preoperative heart failure and its timing, IABP insertion timing, CPB duration and white blood cell count as the leading predictors. The high negative predictive value is what supports the intended use, since it allows most patients to be set aside with confidence so that attention concentrates on the flagged minority.

Against this background, the contributions of this paper are the following.

1. **A multi-modal late fusion pipeline for a registry endorsed composite outcome,** pairing STS ACSD variables with 26 raw intraoperative time series from the same operations to predict a composite of eleven STS National Quality Forum endorsed events in 1,393 consecutive adult cardiac surgery patients, September 2022 to April 2024.
2. **A heterogeneous ensemble matched to the structure of the data,** combining a dual-branch deep network with XGBoost, L1-regularized logistic regression, a support vector machine and a two-stage gradient boosting cascade, with isotonic calibration.
3. **A quantified comparison of the two modalities,** training the identical pipeline on each modality and on both under one cross-validation scheme, which separates what the intraoperative record adds from what the registry already provides.
4. **A per-signal ablation of the intraoperative record,** withholding each time series variable in turn and reporting the resulting changes in AUC, sensitivity and specificity, which separates signals carrying non-redundant information from those overlapping with correlated channels.
5. **An evaluation framed around the intended clinical use,** reporting three named operating points and calibration error alongside discrimination, and interpreting sensitivity, PPV and NPV in terms of screening at ICU handoff.

Consistent with this framing, the novelty of the study lies not in the learning algorithms, which are established, but in two findings that the design makes visible and that the studies reviewed above are not structured to produce. The first is empirical: because the identical pipeline is trained on each modality separately and on both, we can report that the intraoperative signal set does not add discriminative power beyond the registry variables in this cohort, a comparison that prior work adding intraoperative data has not published. The second is methodological: because every signal is ablated in turn, we can show that single-feature ablation on a correlated sensor array measures substitutability rather than importance, which bears on how such ablation results should be interpreted generally. A negative result of the first kind is informative for any group considering the infrastructure cost of waveform capture, and the second qualifies a technique in common use.

## 2 Methods

### 2.1 Data Source

Preoperative patient characteristics and procedural variables from the STS ACSD [5] were paired with detailed time series intraoperative data for adult patients who underwent cardiac surgery with cardiopulmonary bypass at MaineHealth Maine Medical Center over a period from September 2022 to April 2024. 1,393 patients were included. All personal identifiers and private health information (PHI) were removed to protect patient confidentiality. Data was managed according to the ethical standards for medical research involving human subjects, ensuring the reproducibility and portability of our research across other institutions using similar datasets.

### 2.2 Cohort

The cohort for this study was defined to identify patients likely to develop one or more of eleven specific adverse events post-cardiac surgery, as endorsed by STS National Quality Forum (NQF) measures. These adverse events are listed in Table 2.

From the 1,393-patient dataset from the MMC STS registry, we identified 161 patients who developed at least one adverse event. This group formed our case set, as shown in Table 2. The control set comprised remaining patients from the dataset who did not develop any of the adverse events.

### 2.3 Preprocessing and Rationale for Model Selection

In this study we developed a multi-modal late fusion approach [2] for predicting adverse events following cardiothoracic surgery. The dataset comprised two distinct modalities: static patient features from the STS ACSD, and intraoperative time series from physiological parameters collected during cardiac procedures.

For preprocessing, we applied transformations based on feature type. Ratio features received log transformations to preserve their mathematical properties, and the remaining features underwent power transformation to approximate normality; both steps matter because the linear and margin based component models below assume comparable scales and are sensitive to skew, whereas the tree based models are not. Time series data for deep learning was padded to uniform length, since the operations differ in duration and the convolutional and recurrent layers require a fixed input length, and scaled using robust methods, which limits the influence of artifact spikes and sensor dropouts.

No single model family satisfies all the requirements of this task, which is why an ensemble was used. Four properties of the data drove the selection: the modalities differ in structure, so representations must be learned separately before combining, which motivates late fusion over early concatenation; the static modality is a table of mixed continuous, binary and ordinal variables with non-linear interactions and occasional missingness, the regime where boosted trees are effective; the signals contain both transient events and slow trends across an operation lasting hours, calling for a convolutional receptive field for the former and a recurrent state for the latter; and only 161 positive cases are available, which penalizes any single high-capacity model and favours averaging over learners whose inductive biases differ. Table 3 states the mapping explicitly.

**Table 3:**
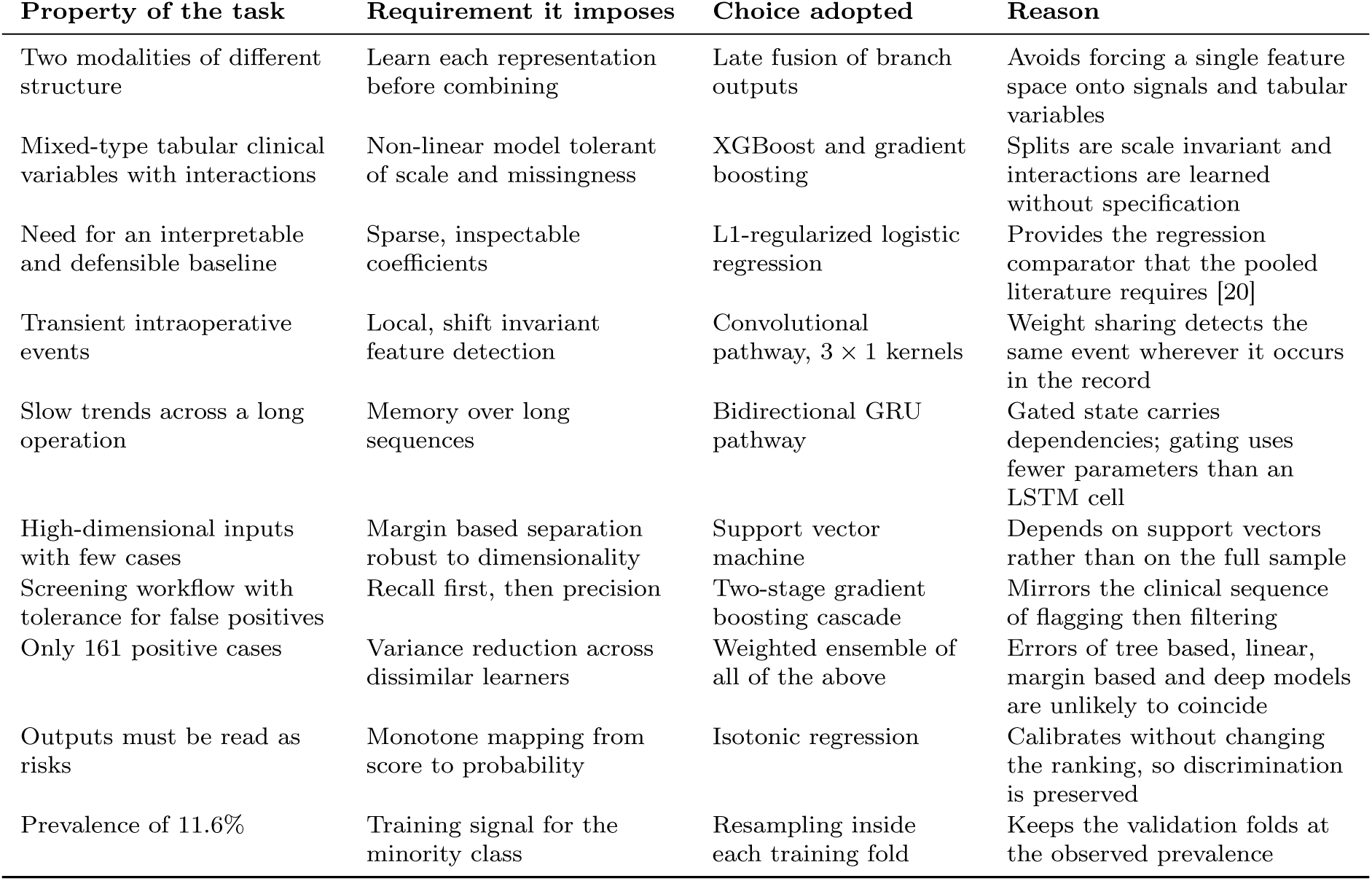
Mapping from properties of the prediction task to the modeling choices adopted in this study.

| Property of the task | Requirement it imposes | Choice adopted | Reason |
| --- | --- | --- | --- |
| Two modalities of different structure | Learn each representation before combining | Late fusion of branch outputs | Avoids forcing a single feature space onto signals and tabular variables |
| Mixed-type tabular clinical variables with interactions | Non-linear model tolerant of scale and missingness | XGBoost and gradient boosting | Splits are scale invariant and interactions are learned without specification |
| Need for an interpretable and defensible baseline | Sparse, inspectable coefficients | L1-regularized logistic regression | Provides the regression comparator that the pooled literature requires [20] |
| Transient intraoperative events | Local, shift invariant feature detection | Convolutional pathway, $3 \times 1$ kernels | Weight sharing detects the same event wherever it occurs in the record |
| Slow trends across a long operation | Memory over long sequences | Bidirectional GRU pathway | Gated state carries dependencies; gating uses fewer parameters than an LSTM cell |
| High-dimensional inputs with few cases | Margin based separation robust to dimensionality | Support vector machine | Depends on support vectors rather than on the full sample |
| Screening workflow with tolerance for false positives | Recall first, then precision | Two-stage gradient boosting cascade | Mirrors the clinical sequence of flagging then filtering |
| Only 161 positive cases | Variance reduction across dissimilar learners | Weighted ensemble of all of the above | Errors of tree based, linear, margin based and deep models are unlikely to coincide |
| Outputs must be read as risks | Monotone mapping from score to probability | Isotonic regression | Calibrates without changing the ranking, so discrimination is preserved |
| Prevalence of 11.6% | Training signal for the minority class | Resampling inside each training fold | Keeps the validation folds at the observed prevalence |

### 2.4 Component Models

#### Dual-branch deep network

The deep learning component, shown in Figure 1, has two branches. The static features branch is a deep residual neural network with three residual blocks of decreasing dimensionality (512 *→* 256 *→* 128), each incorporating batch normalization, dropout and LeakyReLU activations. Skip connections facilitate gradient flow and matter here because a plain stack of this depth trained on a small sample tends to degrade with added layers, whereas a residual block can represent the identity mapping and so never performs worse than its input; batch normalization, dropout and LeakyReLU regularize and keep gradients alive at this case count.

The time series branch featured a parallel structure with two pathways. The convolutional pathway applies 3 *×* 1 kernels along the time axis, so a kernel learns one local waveform motif and applies it at every time point. This weight sharing lets a transient event such as an abrupt pressure fall be detected wherever it occurs, far more efficiently than a fully connected layer over the same window, but the receptive field is bounded by kernel size and depth, which is why it is paired with the second pathway. The bidirectional GRU pathway uses a hidden size of 64 in each direction, giving 128 units once the directions are concatenated, and maintains a gated recurrent state whose update and reset gates let dependencies persist across many time steps without the vanishing gradients of an ungated recurrence. The GRU was chosen over an LSTM for its smaller parameter count at this sample size, and the bidirectional configuration is legitimate because prediction is made retrospectively over a completed operation, so both earlier and later context are available. The two pathways are merged into a dense layer of width 64 with dropout 0.3. The static and time series branches were then combined through late fusion by concatenation, followed by dense layers of width 128, 64 and 32 with dropout rates of 0.3, 0.3 and 0.2 respectively, and a sigmoid output unit.

The network was trained for 50 epochs with a batch size of 32 using the Adam optimizer at a learning rate of 0.001 with gradient norm clipping at 1.0, minimizing binary cross-entropy with class weights of 1.0 for the majority class and min(5.0*, n_−_/n*_+_) for the minority class. Two callbacks were used: the learning rate was halved after eight epochs without improvement in validation loss, down to a floor of 10*^−^*^5^, and training was stopped with a patience of 15 epochs against an equally weighted average of sensitivity and positive predictive value, with the best weights restored. Monitoring that composite rather than loss or accuracy reflects the screening objective, since at an event rate of 11.6% a model can achieve low loss while being clinically useless.

#### XGBoost

XGBoost fits an additive ensemble of regression trees in stages, each fitted to the gradient of the loss and added with shrinkage under an objective that penalizes tree complexity. It was included because its splits are invariant to monotone rescaling, it learns interactions without their being specified, it handles missing values by learning a default branch direction, and it yields feature attributions reportable to clinicians. It was configured with hyperparameters balancing overall performance with calibrated probability outputs, listed in Table 4, including a deliberately small learning rate with a large number of shallow trees, feature and row subsampling, and a positive class weight that compensates for the event rate. Its weakness in this setting is that it has no notion of temporal order, so on the time series modality it operates on features extracted from the signals rather than on the sequences themselves. The feature importance rankings reported in Section 4 are taken from this XGBoost component; they therefore describe which inputs that model relies on, and should be read as indicative of the ensemble rather than as an attribution for it, since the other component models weigh the same inputs differently.

**Table 4:** Hyperparameter settings of the component models, selected by cross-validated grid search and then held fixed.

| Model | Settings |
| --- | --- |
| XGBoost | 500 trees, maximum depth 5, learning rate 0.01, subsample 0.8, column subsample per tree 0.8, minimum child weight 3, $\gamma = 0.1$, $\alpha = 0.01$, $\lambda = 1.0$, positive class weight 5.0, logistic objective, histogram tree method |
| Two-stage boosting, stage 1 (recall) | 300 trees, maximum depth 5, learning rate 0.05, subsample 0.8, minimum samples per split 10, maximum features 0.8; threshold lowered from 0.3 in steps of 0.02 until training recall reaches 0.9 |
| Two-stage boosting, stage 2 (precision) | 300 trees, maximum depth 3, learning rate 0.01, subsample 0.7, minimum samples per split 20, maximum features 0.7; trained on stage 1 positives only, requiring at least 50 samples with more than 10 per class; final score $0.3 \times \text{stage 1} + 0.7 \times \text{stage 2}$ |
| Logistic regression | L1 penalty, $C = 0.05$ ($\lambda = 20$), liblinear solver, balanced class weights, up to 2000 iterations |
| Support vector machine | Radial basis function kernel, $C = 1.0$, scale heuristic for $\gamma$, balanced class weights, probability outputs enabled |
| Deep network, static branch | Dense 512, then residual blocks of width 512, 256 and 128 with batch normalization, LeakyReLU ($\alpha = 0.1$) and dropout 0.3, then dense 64 |
| Deep network, time series branch | Masking, then in parallel a Conv1D layer with 64 filters of length 3 and same padding, ReLU, batch normalization and global average pooling, and a bidirectional GRU with hidden size 64 per direction; merged into dense 64 with dropout 0.3 |
| Deep network, fusion head | Dense 128, 64 and 32 with dropout 0.3, 0.3 and 0.2, sigmoid output |
| Deep network, training | 50 epochs, batch size 32, Adam at learning rate 0.001 with gradient clipping at norm 1.0, binary cross-entropy, minority class weight $\min(5.0, n_-/n_+)$, learning rate halved after 8 stagnant epochs to a floor of $10^{-5}$, patience 15 on the mean of sensitivity and PPV with best weights restored |
| Resampling | SMOTEENN at sampling strategy 0.6, falling back to borderline SMOTE and then plain SMOTE at the same ratio; synthetic sequences formed as Dirichlet-weighted averages of up to three positive sequences with Gaussian noise at 2% of each channel's standard deviation |
| Ensemble | Weighted average over six candidate weight vectors: uniform, and five vectors assigning 0.6 to one model and 0.1 to the rest, scored by the mean of sensitivity and PPV |
| Validation | Stratified 5-fold cross-validation with shuffling, seed 42; inner 80/20 training and monitoring split with seed $10k$ in fold $k$ |

#### L1-regularized logistic regression

This model represents the log odds of an adverse event as a linear combination of the features, with an L1 penalty driving uninformative coefficients to zero, yielding a sparse, inspectable model. It was fitted with balanced class weights and strong regularization (*C* = 0.05, equivalently *λ* = 20), which provides interpretable results with controlled complexity, and it serves a second purpose: since pooled evidence shows no significant discrimination advantage for machine learning over logistic regression in this clinical problem [20], including a regularized regression on the same folds makes the margin attributable to the more complex models measurable rather than assumed. Its limitation is that effects are additive on the log-odds scale, which is part of the reason the skew-correcting transformations described above were applied.

#### Support vector machine

The support vector machine separates the classes by the maximum-margin hyperplane with slack for difficult cases, so the boundary depends only on the support vectors near it rather than on the bulk of the sample. This behaviour is attractive when features are numerous relative to cases. Because its raw output is a signed distance from the boundary rather than a probability, it depends on the calibration step described below; as reported in Section 4, it was the weakest single learner here, and its contribution is as a decorrelated voice in the ensemble rather than as a standalone model.

#### Two-stage gradient boosting

This component is a cascade in which the first stage is optimized for recall and the second for precision. The first stage is deliberately permissive: its decision threshold starts at 0.3 and is lowered in steps of 0.02 until training recall reaches 0.9. The second stage is trained only on the cases the first stage flags, and only when that set contains at least 50 samples with more than ten of each class; the final score for a flagged case is a weighted combination of the two stages, with weights of 0.3 and 0.7 respectively. The design mirrors the clinical workflow the model is intended to support, in which a broad flag is followed by closer review, and it concentrates the second stage’s capacity on the region of the input space where the decision is genuinely difficult. Its known risk is error propagation: a case missed by the first stage cannot be recovered by the second.

### 2.5 Fusion, Calibration and Threshold Selection

The calibrated predictions of the five component models, ordered as deep network, logistic regression, XG-Boost, two-stage gradient boosting and support vector machine, were combined as a weighted average. Rather than optimizing the weight vector continuously, the weights were selected per fold from six fixed candidate vectors: uniform weighting of 0.2 for each model, and five vectors assigning 0.6 to a single model and 0.1 to each of the others. Candidates were scored by an equally weighted average of sensitivity and positive predictive value. Restricting the search to a coarse simplex of this kind is a deliberate regularization choice, since with 161 positive cases a finely optimized weight vector would reflect fold composition more than model quality. This approach allowed us to leverage the strengths of each model while compensating for individual weaknesses, and it is justified by the diversity of the component learners: averaging reduces variance to the extent that the errors being averaged are not identical, and tree based, linear, margin based and deep learners fail on different cases.

To ensure robust probability estimates we applied isotonic regression for probability calibration on each model’s predictions. Isotonic regression fits a non-parametric, non-decreasing mapping from raw score to probability, chosen over a parametric alternative because it imposes no functional form on the miscalibration it corrects. Because the mapping is monotone it leaves the ranking of patients unchanged, so calibration alters the reported probabilities and the behaviour at a threshold without altering AUC. The corresponding risk, which we report on in Section 4, is that a flexible monotone fit can overfit the calibration data when the sample is small, and the ensemble’s calibration error is in fact slightly larger than that of its components.

Because the intended use is screening rather than diagnosis, the decision threshold was selected explicitly rather than left at a default value. We evaluated three strategies: PPV-optimal, which takes the highest-precision point on the precision-recall curve that still achieves a sensitivity of at least 0.7, relaxing that floor to 0.6 and then to 0.5 if no point qualifies; F1-optimal, balancing precision and recall; and balanced, equalizing sensitivity and specificity. Reporting all three makes the sensitivity and specificity trade-off visible to a reader who may weigh false alarms and missed events differently from the way we do.

### 2.6 Validation Protocol and Experimental Configurations

We implemented a stratified five-fold cross-validation framework, with shuffling, and resampling to address class imbalance while preserving data quality. Resampling was applied to the training portion of each fold, so that the held-out folds retain the observed event rate and the reported metrics correspond to the prevalence a deployed model would face. Within each training fold, a further stratified split reserved 20% of the data for monitoring the deep network’s training callbacks.

Class balance was addressed with SMOTEENN at a sampling strategy of 0.6, so the minority class was oversampled to 60% of the majority size and then cleaned by edited nearest neighbours, with fallback to borderline SMOTE, plain SMOTE and finally the unresampled data. A cleaning step was preferred because naive interpolation in a clinical feature space readily produces implausible patients. Synthetic time series rows were generated separately as Dirichlet-weighted averages of up to three randomly chosen positive sequences with Gaussian noise at two percent of each channel’s standard deviation, which keeps each trace a convex combination of real operations while avoiding duplicates.

We trained our models in three configurations, on static features only, on time series features only, and on both, and then performed an ablation study removing one time series feature at a time from the combination of both static and time series features. The purpose of the three configurations is attribution rather than model selection: comparing them isolates what the intraoperative record contributes beyond the registry variables, and the ablation resolves that contribution down to individual monitoring channels.

Hyperparameter values for all component models are given in Table 4. Values were selected by cross-validated grid search and then held fixed for the experiments reported here. A random seed of 42 was fixed for the fold assignment, the resampling and every estimator, and the inner training and monitoring split within fold *k* used seed 10*k*. Experiments were run under Python 3.10.16 with TensorFlow 2 on CPU.

## 3 Statistical Analysis

We performed statistical analysis to compare demographic and clinical factors between the control cohort and cases cohort. The analysis was conducted using independent t-tests for continuous variables (Patient Age, Calculated BMI) to compare the means between the control and case cohorts. Chi-square tests were used for categorical variables (Sex, Race/Ethnicity, ASCVD, Cardiovascular Disease, CAD, preoperative Heart Failure, Hypertension, Diabetes, Current Smoker) when expected frequencies were sufficiently large. Fisher’s Exact Test was used for categorical variables when expected frequencies were small (less than 5). Table 5 presents the calculated p-values for each characteristic, indicating the significance of differences observed between the two cohorts.

**Table 5:**
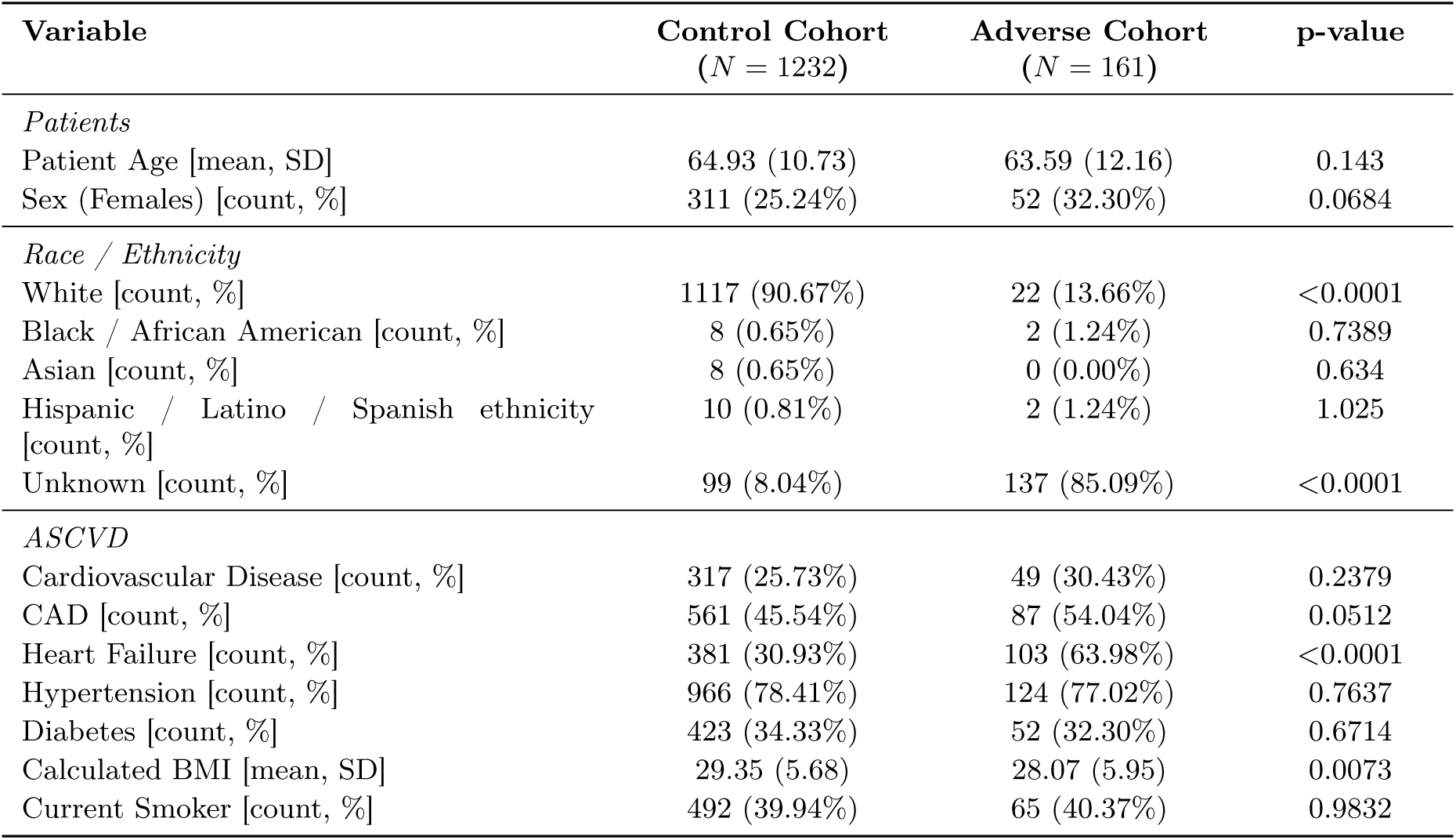
Distributions of characteristics of patients. The race and ethnicity rows reflect differential completeness of the race field between the two cohorts, as discussed in the text, and are not interpreted.

The race and ethnicity rows of Table 5 reflect differences in how completely that field was recorded in the two groups, so we report them for completeness and do not interpret them.

## 4 Results

### 4.1 Predictive Performance of ML Model on both Static and Time Series Data

Our multimodal ensemble model combining static patient characteristics and intraoperative physiological time series demonstrated excellent discriminative ability. As shown in Table 6, the ensemble achieved an AUC of 0.87 (95% CI: 0.82–0.91), outperforming all individual component models, with sensitivity of 0.76, specificity of 0.84, PPV of 0.40 and NPV of 0.97 at the balanced threshold. Gradient boosting ranked second at 0.86 and the SVM lowest at 0.69.

**Table 6:** Model performance summary on both static and time series data.

| Model | AUC | Sensitivity | Specificity | PPV | NPV | F1 Score |
| --- | --- | --- | --- | --- | --- | --- |
| Ensemble | 0.87<br>(0.8166–0.9133) | 75.80%<br>(73.28–78.31) | 83.20%<br>(76.15–90.24) | 40.54%<br>(29.95–51.12) | 96.27%<br>(95.61–96.94) | 51.91%<br>(42.46–61.36) |
| Gradient Boosting | 0.86<br>(0.8020–0.9098) | 73.28%<br>(71.76–74.80) | 81.56%<br>(70.26–92.87) | 40.48%<br>(28.03–52.93) | 95.81%<br>(95.30–96.32) | 50.44%<br>(39.86–61.02) |
| XGBoost | 0.85<br>(0.7986–0.8972) | 77.05%<br>(73.10–80.99) | 78.97%<br>(70.07–87.87) | 35.82%<br>(26.10–45.54) | 96.24%<br>(95.27–97.21) | 47.98%<br>(38.54–57.42) |
| Logistic Regression | 0.78<br>(0.7417–0.8089) | 79.49%<br>(73.82–85.16) | 60.54%<br>(47.61–73.47) | 22.62%<br>(17.58–27.67) | 95.78%<br>(95.10–96.45) | 34.56%<br>(29.12–40.00) |
| Deep Learning | 0.72<br>(0.6778–0.7707) | 87.54%<br>(78.19–96.89) | 36.68%<br>(26.19–47.16) | 15.47%<br>(14.52–16.41) | 96.45%<br>(94.35–98.56) | 26.19%<br>(25.00–27.38) |
| SVM | 0.69<br>(0.6219–0.7688) | 85.00%<br>(77.98–92.02) | 41.04%<br>(22.26–59.83) | 16.78%<br>(14.22–19.35) | 96.37%<br>(94.77–97.98) | 27.77%<br>(24.43–31.12) |

Figure 2 displays the ROC curves for all component models and the ensemble, whose curve dominates across operating points. Feature importance, in Figure 3, shows contributions from both modalities. Heart failure emerged as the most critical predictor, with timing of heart failure onset representing the highest importance feature, followed by timing of intra-aortic balloon pump (IABP) insertion, presence of heart failure at any time point, pre-operative cardiogenic shock and New York Heart Association (NYHA) heart failure classification. Time-based metrics such as duration of cardiopulmonary bypass (CPB) contributed significantly to predictions. Laboratory values including WBC count and last creatinine level, along with vascular comorbidities like peripheral artery disease, further enhanced the model’s predictive capability. Additional factors such as presence of intraoperative PA catheter monitoring, endocarditis risk factors, and valvulopathy provided complementary predictive value. This integration of static clinical factors with dynamic physiological monitoring provided a more comprehensive risk profile than either modality alone.

**Figure 2:**
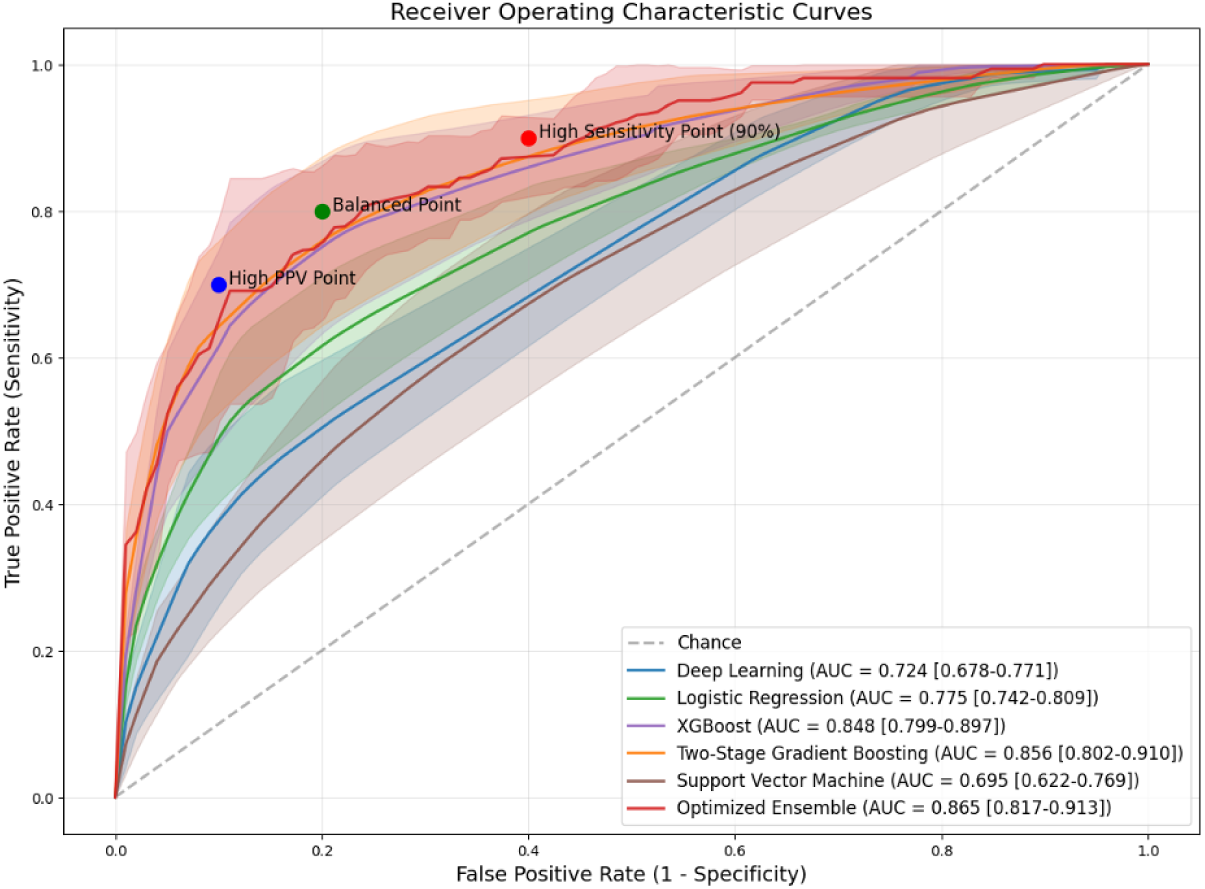
Receiver operating characteristic (ROC) curves for all models used on combined data.

**Figure 3:**
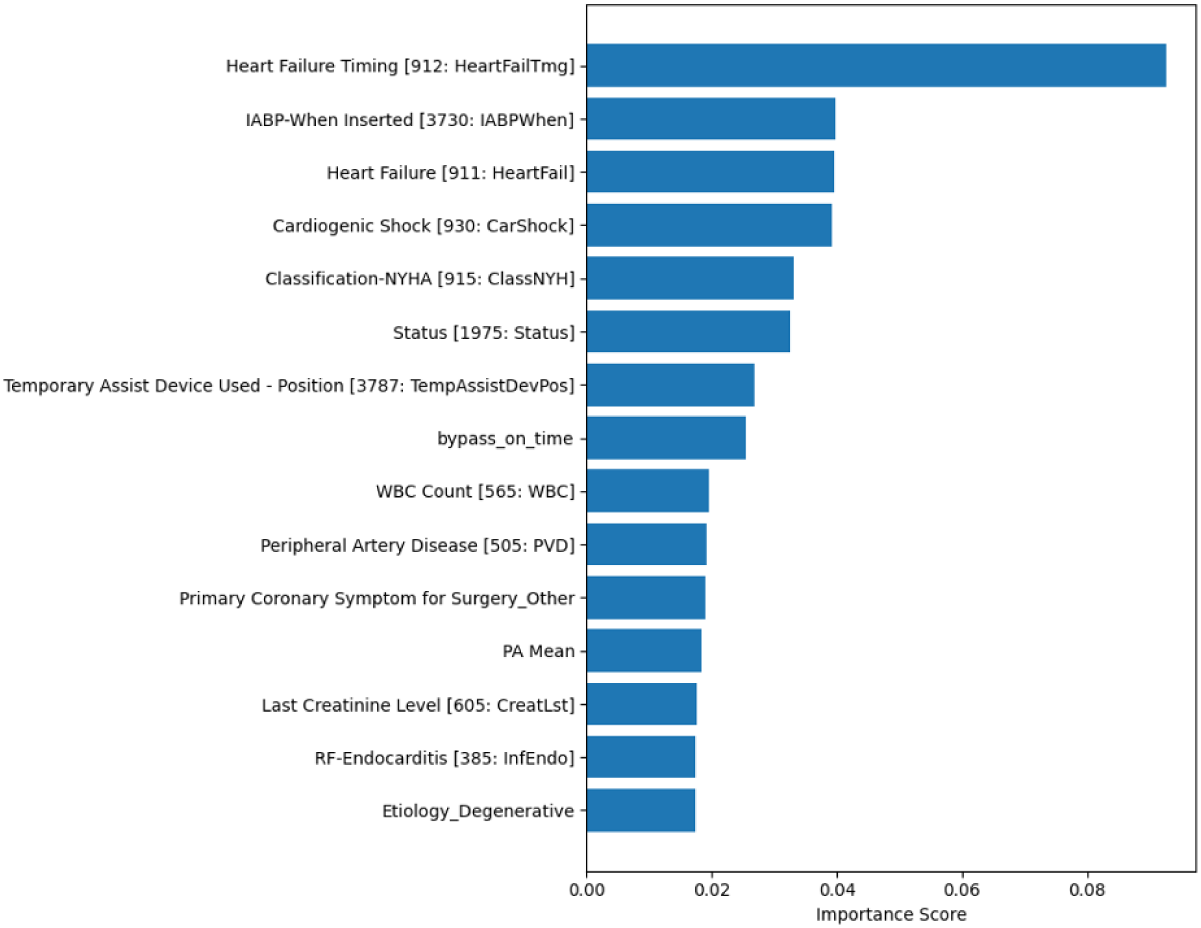
Feature importance on combined data (static and time series), from the XGBoost component model [39].

The calibration plot shown in Figure 4 reveals that while individual component models demonstrate near-perfect calibration with calibration errors close to 0, the ensemble model shows a slight deviation with a calibration error of 0.036.

**Figure 4:**
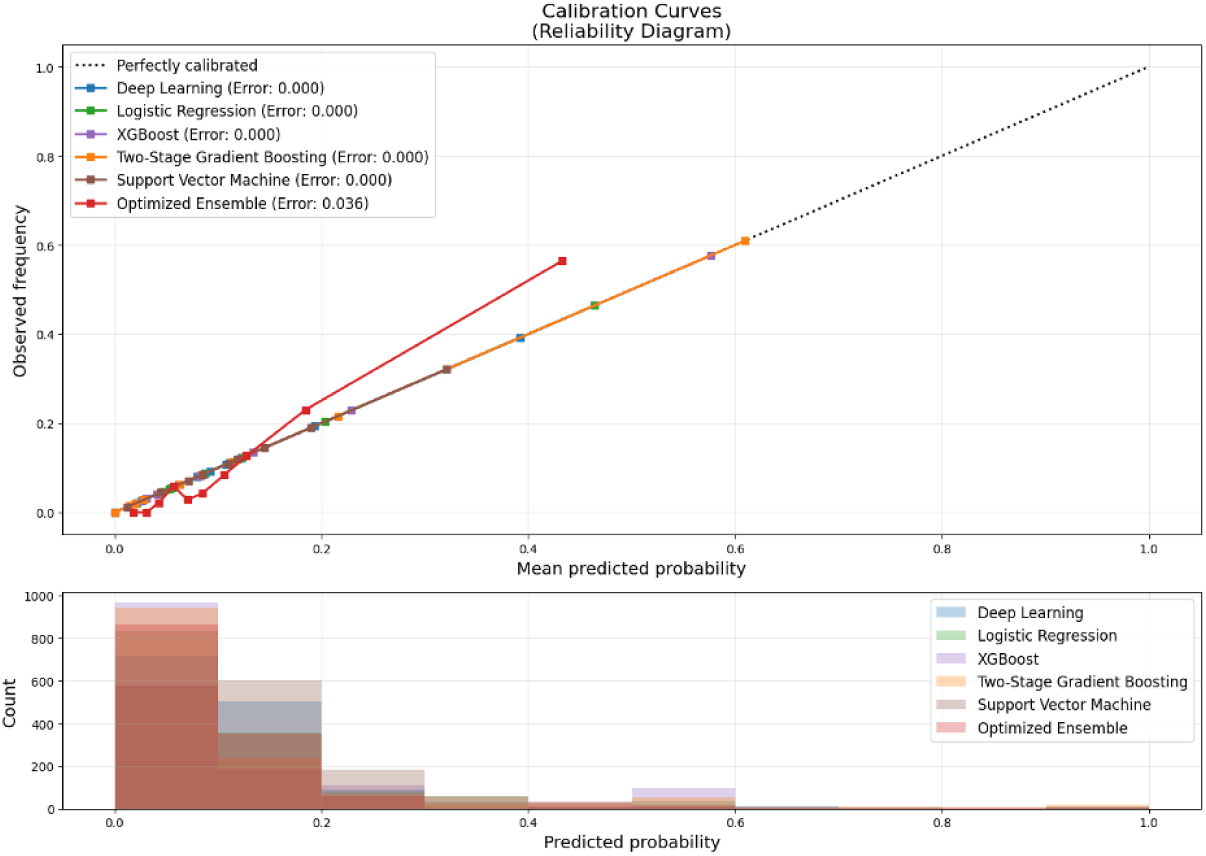
Calibration curve for models on combined data.

#### What Figure 2 shows, and why

The ordering of the curves is itself the finding. The three tree based models occupy the top of the ranking, with the ensemble at 0.87 and gradient boosting and XGBoost at 0.86 and 0.85, while logistic regression reaches 0.78 and the deep network and support vector machine trail at 0.72 and 0.69. Two readings follow. The first is positive: the margin of the ensemble over L1-regularized logistic regression is 0.09 in AUC, which is larger than the difference the pooled literature reports between machine learning and logistic regression for mortality after cardiac surgery [20], so on this task the additional model complexity is doing measurable work rather than merely appearing to. The second is a caution we state explicitly: the ensemble’s advantage over gradient boosting alone is 0.01 AUC and the confidence intervals overlap substantially (0.817 to 0.913 against 0.802 to 0.910), so these results do not establish that the ensemble discriminates better than its strongest single member. What the ensemble buys is not a higher ceiling but a better balance at the chosen operating point, where it attains the highest F1 of any model in Table 6 while holding sensitivity at 0.76. The weakness the figure also exposes is the deep network, at 0.72 with a sensitivity of 0.88 against a specificity of 0.37, a pattern that indicates a model close to flagging everyone rather than one that has learned to discriminate. With 161 positive cases against a network of this capacity, that is the expected failure mode, and it is the reason the deep branch is one weighted voice among five rather than the estimator on its own. The direct route to improvement here is more cases rather than more architecture: the sequence models in the literature that succeed on physiological data are fitted on cohorts an order of magnitude larger [16, 18]. *Lesson learned:* the ensemble is worth its cost here for stability at the operating point rather than for peak discrimination, and a high-capacity sequence model cannot be judged on sensitivity alone when its specificity is this low.

#### What Figure 3 shows, and why

The ranking is dominated by preoperative cardiac status, with heart failure and its timing, IABP insertion timing, cardiogenic shock and NYHA class occupying the top positions, and the strongest intraoperative contributors, CPB duration and the laboratory values, appearing below them. The clinical reading is that the patient who arrives in the operating room already in decompensated heart failure or on mechanical support is the patient most likely to deteriorate afterwards, and that the operation modifies rather than determines that trajectory. This is consistent with the statistical comparison in Table 5, where heart failure is the single largest between-cohort difference (63.98% against 30.93%, *p <* 0.0001). One caveat belongs with this figure: several of these variables are markers of severity recorded because a patient is already unwell, so the figure identifies whom to watch more than what to change. The modifiable signal in the figure is narrower: CPB duration and the hemodynamic channels are influenced by intraoperative conduct, which is where the actionable content lies. Two improvements would strengthen what can be claimed from Figures 3, 6 and 9. An attribution method that applies to the ensemble rather than to one component, such as permutation importance computed on the ensemble output or SHAP values, would remove the mismatch between the model that is evaluated and the model that is explained. Reporting the stability of each rank across the five cross-validation folds would show which positions are reliable, which matters because a ranking estimated from 161 positive cases can reorder substantially between folds, and a clinician shown a top-ten list is entitled to know how firm the ordering is. *Lesson learned:* the model reads severity of preoperative cardiac dysfunction, so its value lies in identifying whom to watch, and any claim about what to change must rest on the smaller set of intraoperatively modifiable variables.

#### What Figure 4 shows, and why

The component models sit almost on the diagonal while the ensemble departs from it by 0.036. The reason is that isotonic regression is applied to each component, so each is calibrated by construction, while a weighted average of calibrated probabilities need not be, since averaging pulls predictions toward the middle of the range. The resulting error of 0.036 is small and leans toward over-calling risk, which is the safer direction for a screening tool. Calibrating the ensemble output as a final step would remove it. *Lesson learned:* calibrating component models does not calibrate their combination, so calibration must be applied at the level at which predictions are reported.

### 4.2 Predictive Performance of ML Model on Static Data

Our ensemble model trained exclusively on static patient characteristics demonstrated strong discriminative ability. As shown in Table 7, the ensemble achieved an AUC of 0.87 (95% CI: 0.82–0.92) with sensitivity of 0.74, specificity of 0.82, PPV of 0.45 and NPV of 0.96 at the PPV-optimized threshold. Gradient boosting followed at 0.86 and the SVM at 0.69.

**Table 7:** Model performance summary on static data only.

| Model | AUC | Sensitivity | Specificity | PPV | NPV | F1 Score |
| --- | --- | --- | --- | --- | --- | --- |
| Ensemble | 0.87 (0.82–0.92) | 73.88%<br>(71.49–76.28) | 82.78%<br>(70.83–94.74) | 44.48%<br>(28.54–60.42) | 95.97%<br>(95.55–96.40) | 53.15%<br>(40.49–65.82) |
| XGBoost | 0.85 (0.80–0.89) | 82.54%<br>(75.43–89.64) | 69.48%<br>(52.16–86.79) | 32.32%<br>(20.49–44.15) | 97.01%<br>(96.32–97.70) | 44.25%<br>(33.15–55.35) |
| Gradient Boosting | 0.86 (0.81–0.90) | 77.61%<br>(72.24–82.99) | 74.82%<br>(60.09–89.55) | 36.50%<br>(22.05–50.96) | 96.24%<br>(95.66–96.82) | 46.82%<br>(34.75–58.89) |
| Logistic Regression | 0.84 (0.78–0.89) | 82.63%<br>(76.08–89.19) | 68.82%<br>(56.12–81.51) | 31.01%<br>(17.68–44.33) | 96.80%<br>(95.66–97.94) | 42.60%<br>(31.73–53.47) |
| SVM | 0.69 (0.62–0.76) | 85.00%<br>(77.98–92.02) | 40.88%<br>(22.17–59.60) | 16.73%<br>(14.19–19.28) | 96.36%<br>(94.75–97.97) | 27.70%<br>(24.38–31.02) |

Figure 5 presents the ROC curves for the static-only models, where the optimized ensemble attains the highest AUC of 0.872 and its curve lies above the others; the high sensitivity, PPV and balanced operating points are annotated.

**Figure 5:**
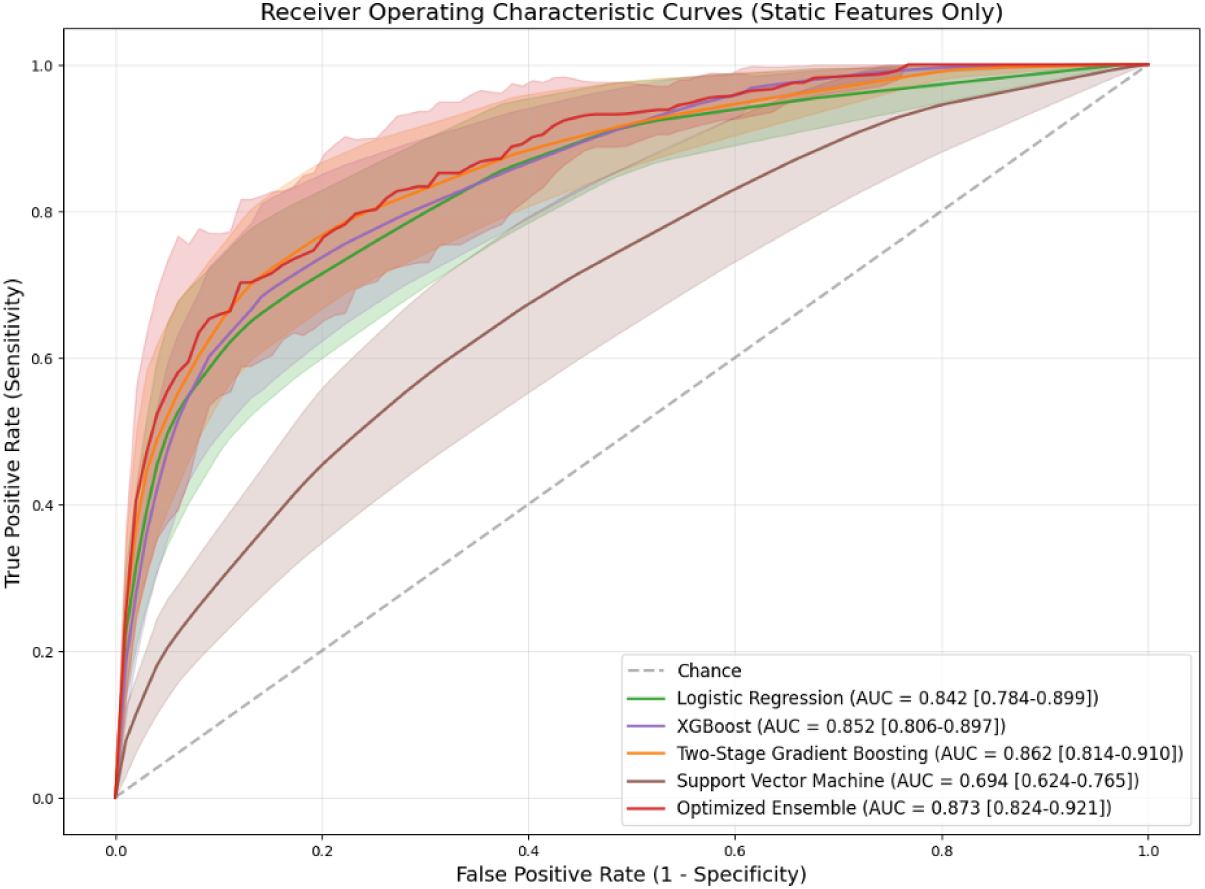
ROC curves for all models used on static data.

Figure 6 shows the top 20 most important features for the static-only ensemble model. The most influential predictor was the presence of heart failure followed by IABP insertion timing and timing of heart failure onset. The presence of cardiogenic shock and urgency of surgical intervention rounded out the top five predictors. Notably, mechanical cardiac support devices featured prominently, with both IABP insertion timing and Temporary Assist Device insertion timing ranking in the top ten features. CPB and aortic cross clamp times demonstrated significant predictive value as did markers of renal function (last creatinine level) and the need to intervene on the tricuspid valve.

**Figure 6:**
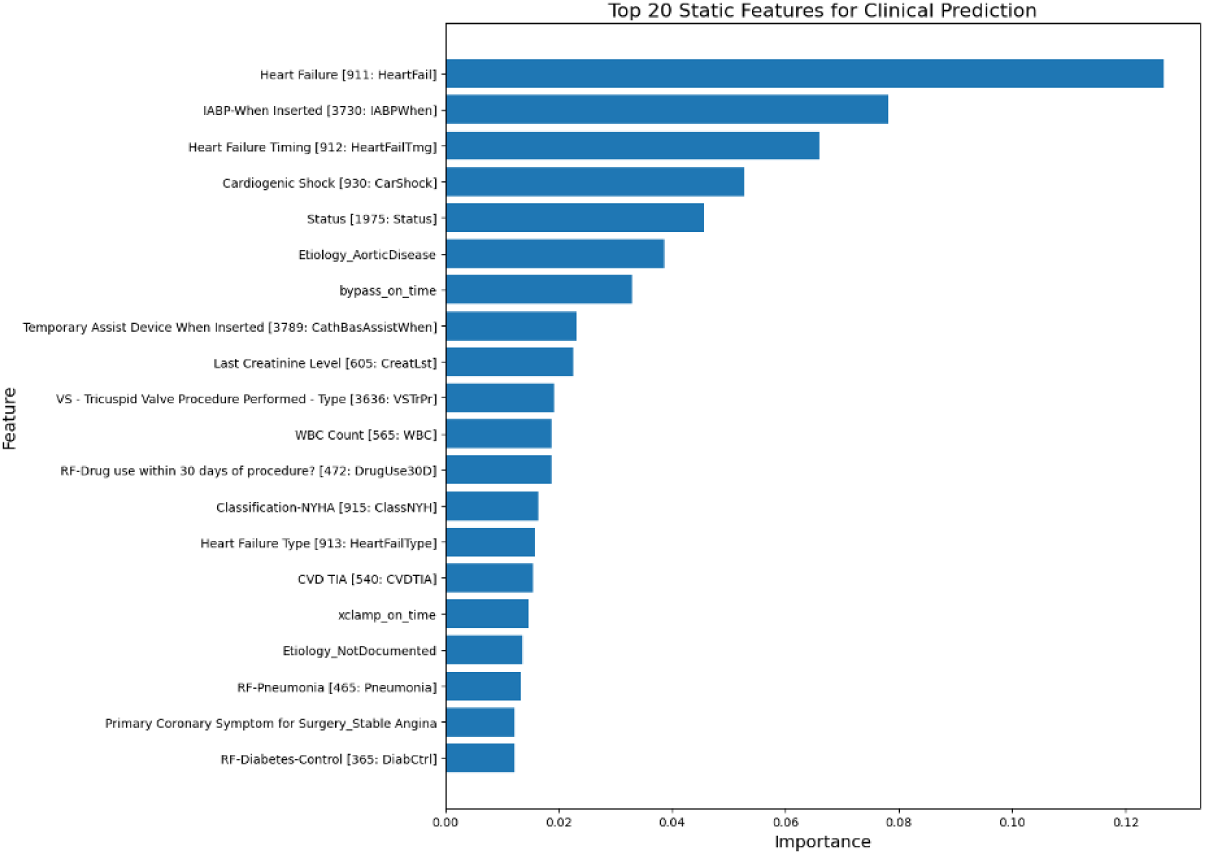
Feature importance on static data, from the XGBoost component model.

Figure 7 gives the calibration curve for the static-only ensemble: the components are near-perfectly calibrated and the ensemble shows a small error of 0.023, visible as slight overestimation in the 0.3 to 0.5 range, a deviation that is clinically tolerable where underestimation is the costlier error.

**Figure 7:**
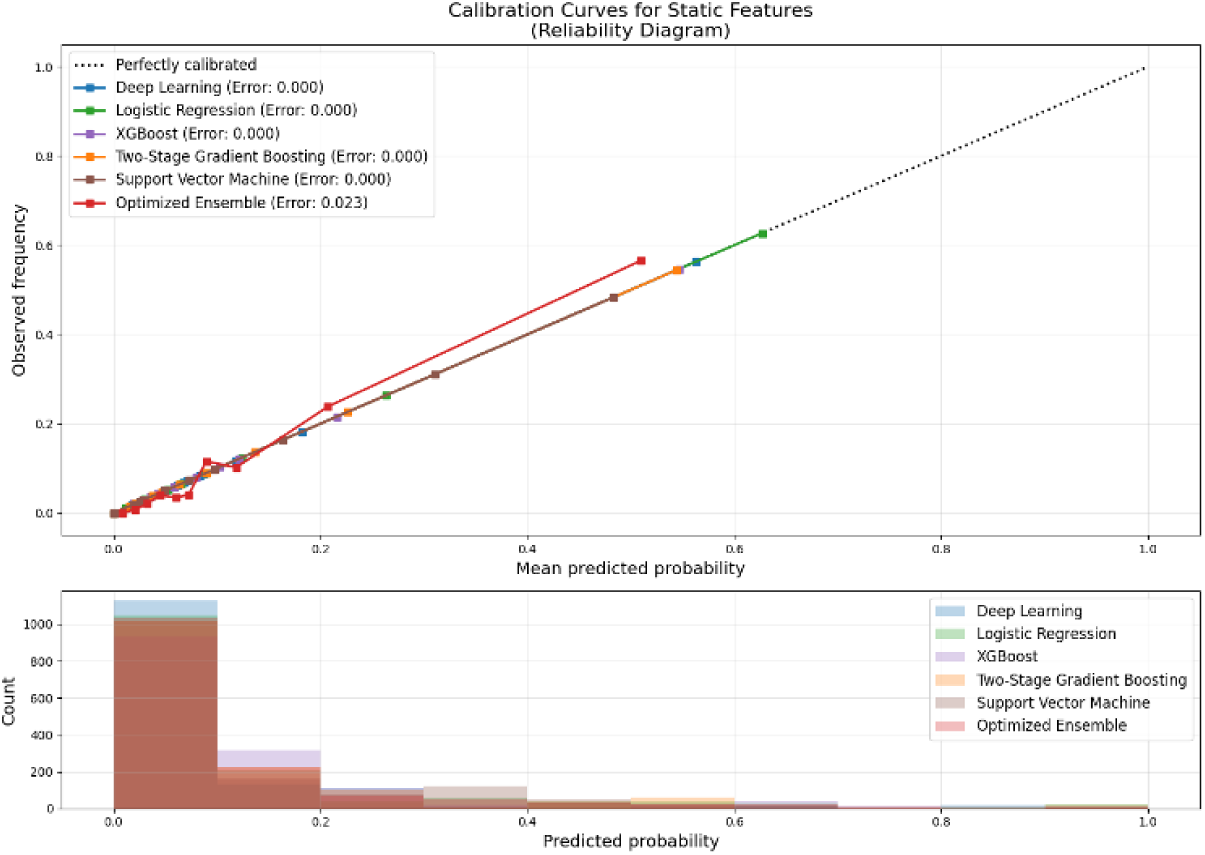
Calibration curve for models on static data.

#### What Figures 5 to 7 show, and why

The most important comparison in this paper is between this subsection and the previous one, and it is not the comparison we expected. The static-only ensemble reaches the same AUC as the combined ensemble, 0.87 against 0.87, and a higher PPV, 0.44 against 0.41. Adding 26 intraoperative signals therefore did not raise discrimination; it shifted the operating characteristics, raising sensitivity from 0.74 to 0.76 and lowering PPV. We report this plainly because the alternative reading, that fusion improved the model, is not supported by Tables 6 and 7. The likely reason is visible in Figure 6 read against Figure 3: the two rankings are close to each other. The variables that dominate the staticsonly model, heart failure presence, IABP insertion timing, heart failure onset timing, cardiogenic shock and urgency of surgery, are the same variables that dominate the combined model, and the intraoperative quantities that matter most in either, CPB and aortic cross clamp duration, are already present in the static record as procedural fields rather than as waveforms. Figure 6 additionally places temporary assist device insertion timing and the need to intervene on the tricuspid valve in its top ten, both of which again describe preoperative cardiac failure and operative complexity rather than intraoperative physiology. The raw signals therefore largely restate severity that the registry has already captured, which is a substantive negative result and one the literature does not report, since prior work added intraoperative data without publishing the preoperative-only comparison on the same folds [28, 29].

A second observation concerns logistic regression. On static data it reaches 0.84, within 0.03 of the ensemble, whereas on combined data it falls to 0.78. A sparse linear model degrades when several hundred correlated signal-derived features are appended to a compact clinical vector, while the tree based models absorb them without loss. This is a concrete instance of the lesson drawn from the pooled literature in Section 1.2: on the registry variables alone, the complex ensemble earns only a small margin over a regularized regression, and a reader entitled to the simplest adequate model would be justified in preferring the regression in a setting where interpretability dominates.

The calibration picture is the same in kind as for the combined model, with the components close to the diagonal and the ensemble at 0.023, a smaller error than the 0.036 of the combined ensemble, consistent with averaging over a lower-dimensional and less heterogeneous input space. The practical conclusion is favourable for deployment: a site with registry data and no access to intraoperative waveforms can expect essentially the discrimination reported here, which considerably lowers the barrier to external validation.

In summary, the static-only ensemble is as discriminative as the full model and easier to deploy, and the case for the intraoperative signals has to rest on what they add at a given operating point and on the attribution they make possible, rather than on AUC. *Lesson learned:* a modality comparison on identical folds can show a new data source to be redundant rather than additive, which is worth reporting precisely because the preoperative-only comparison is usually omitted.

### 4.3 Predictive Performance of ML Model on Time Series Data

Our ensemble model trained exclusively on intraoperative time series data demonstrated good predictive performance, confirming that dynamic physiologic signals offer valuable insight into patient risk profiles. As summarized in Table 8, the ensemble achieved an AUC of 0.83 (95% CI: 0.80–0.86) with sensitivity of 0.75, specificity of 0.78, PPV of 0.33, NPV of 0.96 and an F1 of 0.45. It trails the combined model in discrimination but outperforms every individual learner, which supports its use as a standalone tool in real-time monitoring settings.

**Table 8:**
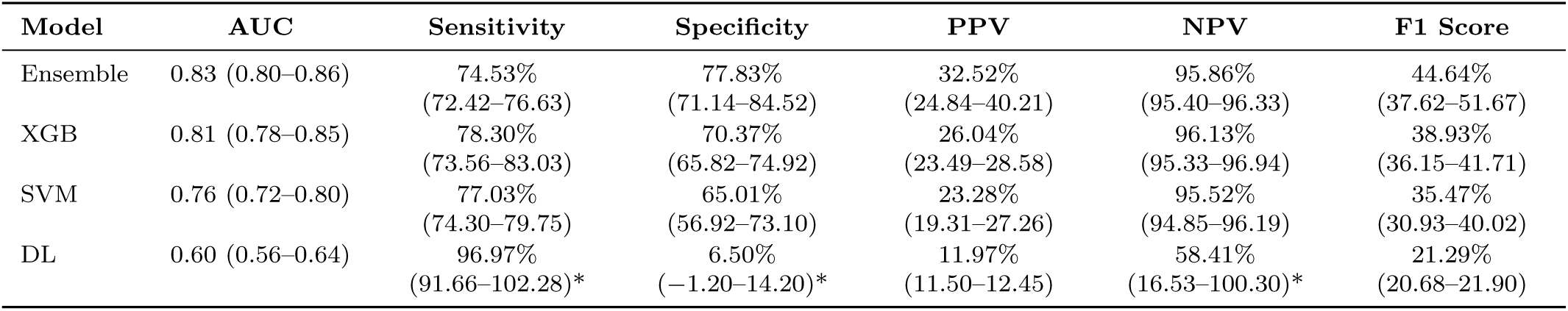
Model performance summary on time series data only. Values marked with an asterisk indicate confidence intervals that extend beyond the feasible range.

Figure 8 presents the ROC curves for the time-series-only models, where the optimized ensemble dominates at an AUC of 0.83 against 0.81 for XGBoost, 0.76 for the SVM and 0.60 for the deep model; the high sensitivity, balanced and high PPV operating points are annotated.

**Figure 8:**
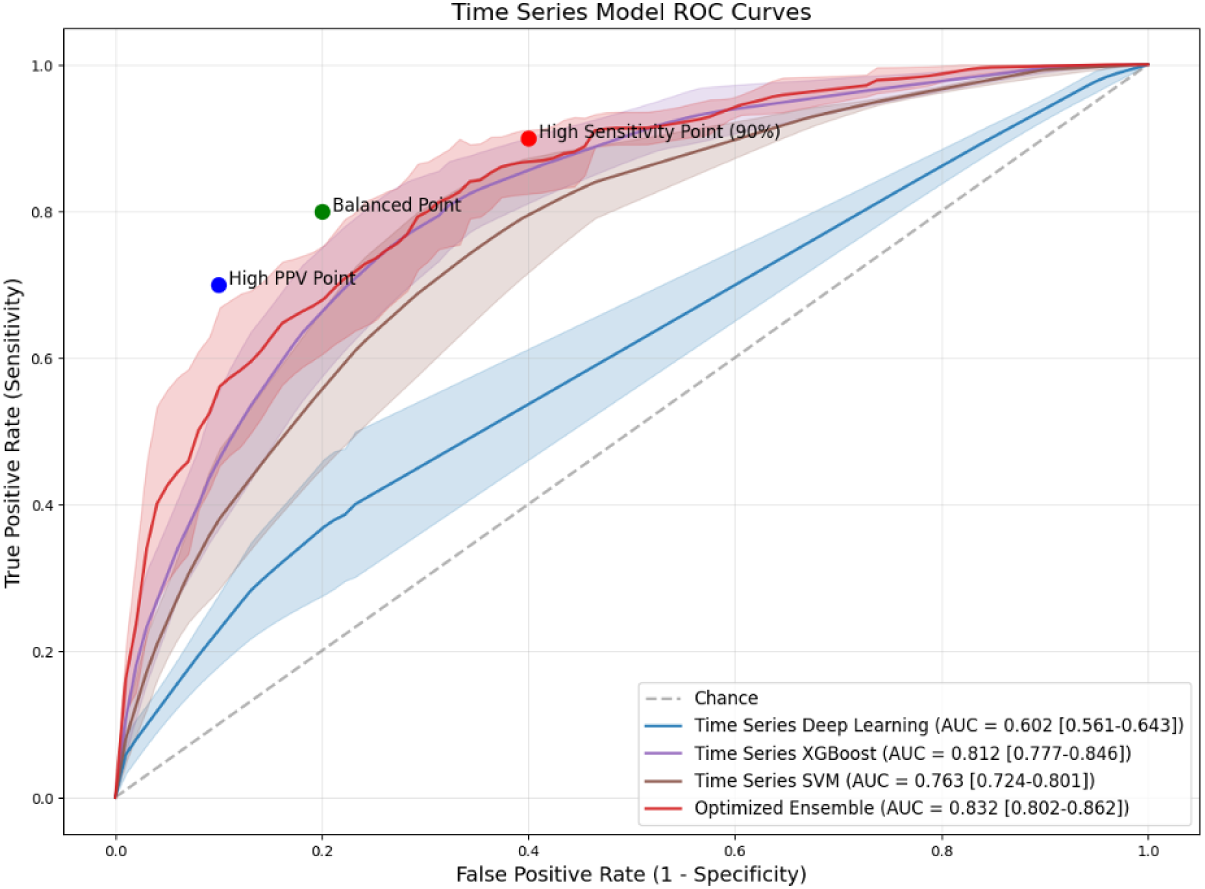
ROC curves for all models used on time series data.

Figure 9 highlights the top 10 time series data-based predictors contributing to the model’s performance. CPB duration emerged as the most important feature, followed closely by vital sign parameters including pulse rate and central venous pressure (CVP). Hemodynamic monitoring proved critical, with arterial flow on CPB ranking highly among predictors. Oxygenation metrics including SpO_2_ and ecDO_2_i (extracorporeal oxygen delivery index) while on CPB demonstrated substantial importance in predicting outcomes. Additional cardiovascular parameters such as ABP mean and pulse pressure, along with aortic cross clamp duration, and core temperature variability further enhanced predictive capability. Taken together, these features illustrate the model’s capacity to integrate hemodynamic stability, oxygenation status, temperature variability and procedural duration to detect patterns that may precede adverse events.

**Figure 9:**
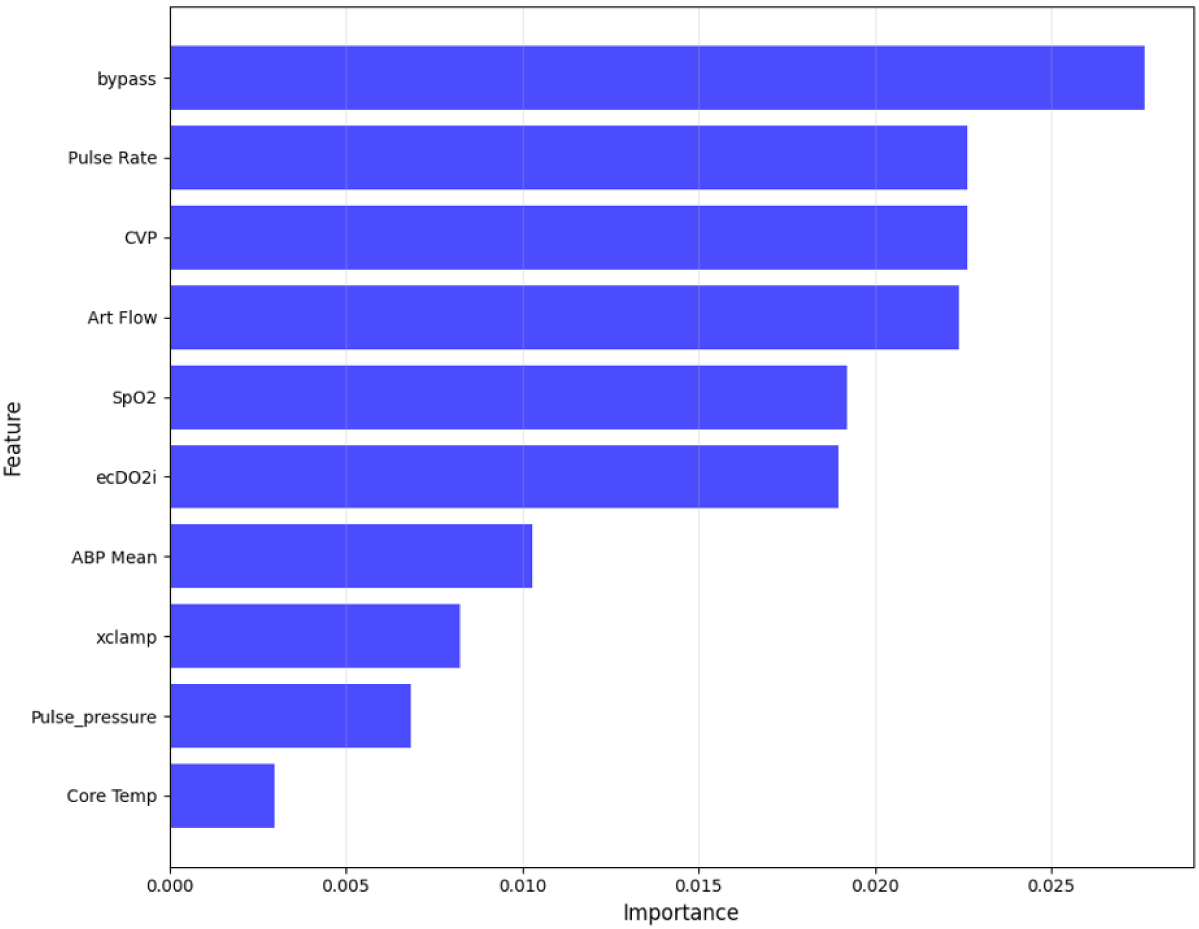
Feature importance on time series data, from the XGBoost component model.

Figure 10 displays the calibration curves. The components are nearly perfectly calibrated and the ensemble deviates by 0.036, again as mid-range overestimation, while otherwise tracking the diagonal closely.

**Figure 10:**
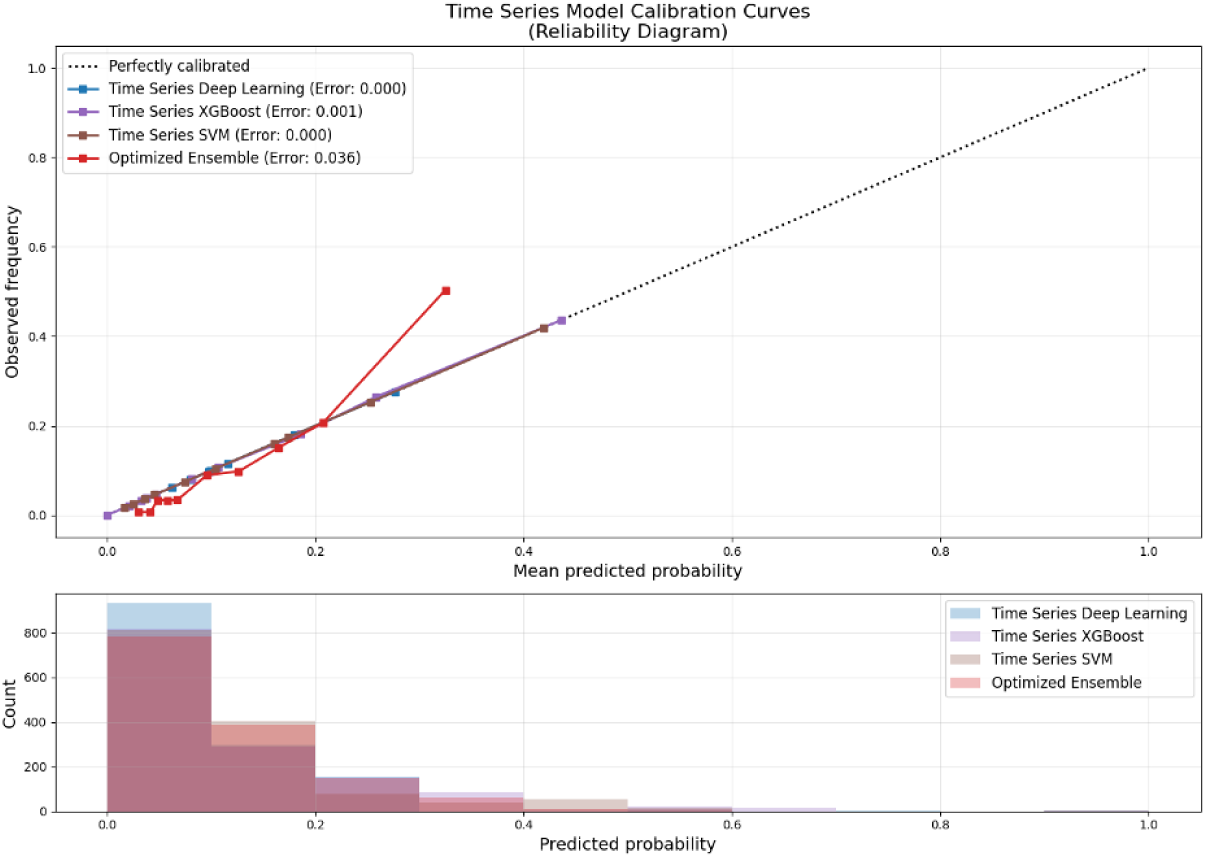
Calibration curve for models on time series data.

Against the static-only model the time series ensemble is slightly more sensitive (0.75 against 0.74) but less specific (0.78 against 0.83), with a lower PPV (0.32 against 0.45), and against the combined model it is lower in both AUC (0.83 against 0.87) and PPV. Its advantage is availability during the operation rather than performance.

#### What Figures 8 to 10 show, and why

The headline of this configuration is that the intraoperative record alone reaches an AUC of 0.83, only 0.04 below the registry variables on their own. For a model that knows nothing about the patient’s cardiac history, comorbidities or the urgency of the operation, that is a substantial result, and it establishes that the signals do carry outcome-relevant information rather than noise. The importance ranking explains where that information sits: CPB duration leads, followed by pulse rate, CVP and arterial flow on bypass, so the model is largely reading duration of physiological support and the adequacy of circulation, which are the same quantities a perfusionist monitors. Read alongside Section 4.2, this shows that each modality reaches most of the achievable discrimination on its own, which means a site can deploy the model with whichever of the two it holds.

This configuration compares most directly with prior work. Fernandes et al. [29] also predicted from intraoperative parameters and located the strongest signal in hypotension occurring outside the bypass phase, that is in arterial pressure behaviour, which is the same family of channels that Figure 9 ranks highly through ABP mean and pulse pressure and that the ablation in Section 4.4 identifies as least substitutable. Two differences matter. That study derived an explicit exposure variable, hypotension burden outside CPB, whereas our pathway learns from the signal directly, and its endpoint was mortality rather than a composite. That two studies converge on arterial pressure from different modelling directions is mutual support for the finding.

The deep network is the exception, at an AUC of 0.60 with sensitivity of 0.97 and specificity of 0.065, which reflects a model labelling almost every patient positive rather than one that discriminates, so we do not treat it as a standalone model on this modality. Learning from the signals alone, with no static context and 161 cases, is the hardest representation problem in the study. Larger cohorts, pretraining the sequence encoder on unlabeled operations, and reporting AUPRC alongside AUC are the natural next steps.

Calibration behaves as in the other two configurations, with the ensemble at 0.036 and the same mid-range overestimation. The consistency of that pattern across all three configurations supports the mechanistic explanation given in Section 4.1, namely that averaging calibrated component outputs compresses them toward the centre, rather than anything specific to a modality.

In conclusion, the time series-only ensemble provides clinical value in settings where registry variables are unavailable at prediction time, and it demonstrates that the intraoperative record is independently informative. Its performance does not exceed that of the registry variables, and its value is therefore best understood as redundancy and availability rather than as additional discriminative power. *Lesson learned:* the intraoperative signals alone carry most of the achievable discrimination, so in this cohort the two modalities act as substitutes rather than complements, which is what makes deployment feasible at sites holding only one of them.

### 4.4 Ablation Study on Time Series Data

We conducted an ablation study to assess the individual contribution of each intraoperative time series variable to the performance of our ensemble model. In this approach, we systematically removed one time series feature at a time while keeping all other features, including static variables and the remaining time series, intact. The model used for this analysis was the same optimized ensemble employed in our main results, with the balanced threshold configuration retained to ensure fair comparison. For each ablation run, we retrained the model using the modified feature set and evaluated it using cross-validation, measuring AUC, sensitivity, and specificity.

Our baseline model, using all available static and time series features, achieved an AUC of 0.870, a sensitivity of 0.76, and a specificity of 0.83. These values served as the reference point for assessing the relative drop or improvement when a particular feature was excluded, as stated in Table 9. Results indicated that no single time series feature was solely responsible for the model’s predictive performance; rather, a combination of features contributed to overall robustness. However, the degree to which performance dropped upon feature removal varied, revealing the relative importance of each variable.

**Table 9:** Ablation study results summary on time series data.

| Feature Removed | AUC | $\Delta$ AUC (%) | Sensitivity | $\Delta$ Sensitivity (%) | Specificity | $\Delta$ Specificity (%) |
| --- | --- | --- | --- | --- | --- | --- |
| Full model (baseline) | 0.87 | — | 0.76 | — | 0.83 | — |
| ecDO<sub>2</sub>i | 0.8789 | 1.08 | 0.7721 | 1.82 | 0.8121 | −2.06 |
| ART Mean | 0.8761 | 0.74 | 0.7111 | −7.36 | 0.8674 | 4.30 |
| AO Mean | 0.8749 | 0.59 | 0.7297 | −4.56 | 0.8471 | 1.97 |
| Pulse Rate | 0.8737 | 0.45 | 0.7291 | −4.65 | 0.8430 | 1.50 |
| ABP Sys | 0.8736 | 0.43 | 0.7431 | −2.54 | 0.8674 | 4.30 |
| ABP Dia | 0.8730 | 0.36 | 0.7479 | −1.82 | 0.8446 | 1.68 |
| ST generic V | 0.8717 | 0.20 | 0.7663 | 0.94 | 0.8219 | −0.93 |
| Esoph Temp | 0.8710 | 0.12 | 0.7663 | 0.94 | 0.8227 | −0.84 |
| AO Sys | 0.8711 | 0.12 | 0.6935 | −10.01 | 0.8845 | 6.27 |
| Art Flow | 0.8710 | 0.11 | 0.7475 | -1.88 | 0.8259 | -0.47 |
| Pulse pressure | 0.8704 | 0.05 | 0.6933 | -10.04 | 0.8788 | 5.61 |
| CI BSA | 0.8705 | 0.05 | 0.6808 | -11.92 | 0.8837 | 6.17 |
| ST generic II | 0.8705 | 0.05 | 0.7538 | -0.94 | 0.8251 | -0.56 |
| ETCO<sub>2</sub> | 0.8704 | 0.04 | 0.7602 | 0.03 | 0.8243 | -0.65 |
| ECG HR | 0.8704 | 0.04 | 0.7055 | -8.21 | 0.8683 | 4.40 |
| ART Sys | 0.8701 | 0.00 | 0.6871 | -10.98 | 0.8820 | 5.99 |
| bypass | 0.8700 | 0.00 | 0.7600 | 0.00 | 0.8300 | 0.00 |
| xclamp | 0.8700 | 0.00 | 0.7600 | 0.00 | 0.8300 | 0.00 |
| Core Temp | 0.8697 | -0.04 | 0.7496 | -1.57 | 0.8560 | 3.00 |
| AO Dia | 0.8691 | -0.11 | 0.7663 | 0.94 | 0.8170 | -1.49 |
| ART Dia | 0.8690 | -0.13 | 0.7602 | 0.03 | 0.8195 | -1.21 |
| SpO<sub>2</sub> | 0.8687 | -0.16 | 0.6810 | -11.89 | 0.8829 | 6.08 |
| CVP | 0.8648 | -0.63 | 0.6873 | -10.95 | 0.8828 | 6.08 |
| ABP Mean | 0.8619 | -0.98 | 0.7229 | -5.59 | 0.8381 | 0.93 |

The exclusion of the ABP Mean signal led to the largest drop in AUC, from 0.870 to 0.862, and removing CVP produced the next largest reduction, to 0.865. Removal of SpO_2_, CVP or ABP Mean lowered sensitivity while raising specificity, by roughly twelve, eleven and six percentage points respectively in the first two cases, which indicates that these channels shift where the decision threshold falls as much as they contribute raw discriminative information.

#### What Table 9 shows, and why

The dominant pattern in the table is not degradation but insensitivity. Every single-signal removal changes AUC by less than 1.1% in either direction, and for the majority of the 24 signals the AUC after removal is marginally higher than the baseline of 0.870. This is a sign of redundancy among the channels. The intraoperative channels are strongly correlated with one another by construction: systolic, diastolic and mean pressures are derived from the same arterial waveform, ART, AO and ABP measure pressure at different sites in the same circulation, and pulse rate and ECG heart rate report the same physiological quantity through different sensors. Removing any one member of such a group leaves the information available through its neighbours, while slightly reducing the dimensionality the model must fit at 161 cases, which is enough to produce the small positive deltas observed. Two signals behave differently, ABP Mean and CVP, and both are the least substitutable members of their groups, mean arterial pressure summarizing perfusion across the cycle and central venous pressure measuring filling rather than ejection. The finding for CVP is consistent with the independent association between elevated CVP and mortality and renal failure after cardiac surgery reported by Williams et al. [41].

The result also carries a methodological point. In a correlated sensor array, withholding one channel measures what that channel adds once every substitute is still present, which is near zero whenever a substitute exists. Grouped ablation, withholding all pressure channels or all temperature channels together, would separate the value of a physical quantity from the value of one sensor reporting it, and adding signals one at a time to the static model would show how quickly the achievable performance is reached. Both run on the existing pipeline without additional data.

The practical implication is favourable for deployment: no single intraoperative channel is load bearing, so a monitoring gap in any one channel during an operation would not invalidate the prediction. Identifying a minimal sensor set would require the grouped analysis described above. *Lesson learned:* in a correlated sensor array, single-feature ablation measures substitutability rather than importance, and only grouped ablation can establish which physical quantity the model depends on.

### 4.5 Comparison with Reported Results in the Literature

Placing these numbers beside published counterparts requires care, because the endpoint, the cohort size and the event rate all differ, and each of those moves the metrics. With that caveat, three comparisons are informative. Against the field-level benchmark, the Bayesian meta-analysis of 51 studies by Penny-Dimri et al. [20] pooled a C-index of 0.82 (95% credible interval 0.79 to 0.85) for machine learning models predicting 30-day mortality after cardiac surgery and 0.81 for in-hospital mortality. Our AUC of 0.87 sits above that interval, but the endpoint differs: a composite of eleven events occurring in 11.6% of patients is a different and in several respects easier prediction target than mortality alone, so we read this as consistent with the literature rather than as evidence of superiority. Against the closest single study, Ferrando-Vivas et al. [28] reported a c-index of 0.90 using pre- and intra-operative data from national critical care units, which exceeds our figure on a far larger and multi-centre sample; our contribution relative to that work is the attribution of performance to modality and to individual signals. Against Bertsimas et al. [21], whose optimal classification trees reached a test AUC of 86.2% on 295,000 congenital operations, our discrimination is comparable on a cohort three orders of magnitude smaller, which is the expected consequence of a narrower and more homogeneous population rather than of a better model.

Two comparisons are deliberately not made. The positive predictive value of 97% reported by Ebel [27] and the observed-to-expected ratio of 0.985 reported by Kilic et al. [26] are not commensurable with our PPV of 0.40, because PPV depends on prevalence and the observed-to-expected ratio measures calibration in the large rather than discrimination. Stating this explicitly matters more than it may appear: a reader comparing headline figures across these studies would conclude that our model is far weaker, when the difference is arithmetic rather than substantive.

Finally, on the question the pooled literature poses most directly, whether machine learning earns its complexity over logistic regression [20], our answer is conditional. On the combined modality the margin is 0.09 AUC in favour of the ensemble, which is larger than the pooled difference; on the static modality alone it narrows to 0.03, which is within the range where the simpler model would reasonably be preferred. The honest summary is that the ensemble earns its complexity when the intraoperative signals are present and earns considerably less of it when they are not.

## 5 Discussion

Our investigation employed an ensemble approach for predicting adverse events after cardiothoracic surgery in 1,393 patients from the STS ACSD at MaineHealth Maine Medical Center between September 2022 and April 2024 [5], achieving an AUC of 0.87 with sensitivity of 0.76 and specificity of 0.83 at the balanced threshold, comparable to the pooled discrimination reported for this clinical problem [20] on a considerably smaller cohort.

The relationship between the two modalities proved more complicated than anticipated. The static-only and combined models achieved the same AUC of 0.87, with the static-only model attaining the higher PPV, while the time series-only model reached 0.83. The intraoperative signals therefore did not add discriminative power on this cohort; they changed the operating characteristic, raising sensitivity from 0.74 to 0.76 at the cost of precision. The ablation study supports reading the modalities as overlapping rather than complementary, since no individual signal proved load bearing. Within that picture, the prominence of hemodynamic parameters, particularly CVP, ABP mean and pulse pressure, is where the clinically modifiable content lies, and elevated CVP has been identified as an independent predictor of mortality and renal failure after cardiac surgery, with each 5 mmHg increase associated with significantly increased risk [41]. The ensemble calibration error of 0.036 leans toward risk overestimation, the safer direction for a screening tool, and calibrating the ensemble output directly would reduce it further.

The contribution of this study lies in its evaluation design rather than in the learning algorithms, which are established and used in standard form. Prior work adding intraoperative information to preoperative models reports the combined result [28, 29] without the single-modality comparison on the same folds, so the incremental value of the added data is not separable from the value of the model. Running all three configurations under one cross-validation scheme and then ablating each signal yields two results a single combined model cannot: that the modalities are substitutes in this cohort, and that per-signal ablation in a correlated array quantifies substitutability rather than importance. Both are negative or qualifying findings, and both are more useful to a group planning such a system than another report of a high AUC. Kurlansky et al. [9], whose work acted as our reference point, addressed a similar purpose on a different dataset; relative to that and to traditional preoperative-only risk models, our contribution is the attribution of performance to modality and to individual signals rather than a higher headline figure.

## 6 Limitations

Despite the promising results, our study has several limitations that should be acknowledged. First, the single-center dataset size is moderate, particularly for the adverse event cases with just 161 patients, which limits the model’s ability to generalize to diverse patient populations. While our cross-validation approach helps mitigate this concern, external validation on larger, multi-center cohorts would strengthen confidence in the model’s applicability.

Second, while our PPV values (ranging from 0.30 to 0.51 across different model configurations) represent an improvement over many existing clinical risk scores, there remains room for enhancement to further reduce false positive predictions. This is particularly important in resource-constrained clinical environments where false alarms can lead to intervention fatigue and resource misallocation.

Third, the feature importance rankings come from the XGBoost component, so they describe that model’s inputs rather than an attribution for the full ensemble; applying a model-agnostic method to the ensemble output is left to future work.

Finally, our model treats diverse adverse events as a single outcome category. Future work with larger datasets could potentially allow the development of models designed to predict a specific adverse event, allowing for more tailored risk prediction and intervention strategies.

## 7 Conclusions and Future Work

### 7.1 Lessons Learned

In this study we developed an ensemble modeling approach for predicting adverse events following cardiothoracic surgery by integrating static patient characteristics with intraoperative physiological measurements, and evaluated it in three modality configurations with a per-signal ablation. The model achieved an AUC of 0.87 with sensitivity of 0.76, specificity of 0.83 and a negative predictive value of 0.96 at the balanced threshold. We set out below what the study taught us, since these lessons transfer beyond this cohort.

1. **The registry variables already contain most of the separable signal.** The static-only configuration matched the combined configuration in AUC, at 0.87, and exceeded it in positive predictive value, 0.44 against 0.41. The practical consequence is favourable: a prediction of this quality is attainable from data that every STS participating site already collects, without waveform infrastructure, which lowers the barrier to external validation and deployment.
2. **The intraoperative record is independently informative but largely substitutable.** Trained alone, the signals reached an AUC of 0.83 without any knowledge of the patient’s history, which establishes that they carry genuine outcome information. That the two modalities together do not exceed either alone indicates that they encode overlapping severity rather than complementary aspects of risk. For this cohort, the modalities behave as substitutes, which matters operationally: a site that has only one of them is not thereby excluded.
3. **No single monitoring channel is load bearing.** Across 24 single-signal ablations, AUC moved by less than 1.1% in either direction, and for most channels removal marginally improved it. Only ABP mean and CVP produced meaningful reductions, and both are the least substitutable members of their correlated groups. The methodological lesson is that single-feature ablation on a correlated sensor array measures substitutability rather than importance, and the clinical one is that a monitoring gap in any one channel would not invalidate a prediction.
4. **The ensemble earned its complexity where the data was richest.** It exceeded L1-regularized logistic regression by 0.09 AUC on the combined modality, a larger margin than the pooled literature reports for machine learning over logistic regression in this clinical problem [20], and by 0.03 on the static modality alone. Reporting both margins lets a reader judge where the added complexity is worth its cost.
5. **Deep sequence models need larger cohorts than this.** The deep network reached an AUC of 0.72 on combined data and 0.60 on signals alone. With 161 positive cases, treating such models as one weighted voice in an ensemble rather than as the primary estimator is the safer design, and it is what allowed the ensemble to benefit from them without being held back by them.
6. **Calibration must be applied where predictions are reported.** The individual models are well calibrated and the ensemble that averages them is not, at an error of 0.036 with risk overestimated in the 0.3 to 0.5 band. Averaging calibrated probabilities does not preserve calibration, which is a general point that applies to any weighted ensemble reporting risks to clinicians.
7. **The operating point, not the AUC, determines clinical value.** The negative predictive value of 0.96 to 0.97 across configurations is what makes the model usable for screening, since most patients can be set aside with confidence. At a PPV of about 0.40, whether the resulting review burden is acceptable is a clinical judgement, which is why we report three named operating points rather than one.

Feature importance across all three configurations pointed consistently to preoperative cardiac status, with heart failure and its timing, IABP insertion timing and cardiogenic shock dominating, while the signal-derived predictors that mattered were CPB duration, pulse rate, CVP and arterial flow on bypass. Taken together these indicate that preoperative cardiac status and the timing of mechanical support are the strongest determinants of adverse events in this cohort, and that the intraoperative contribution is concentrated in duration of support and adequacy of circulation.

### 7.2 Future Work

The most immediate work is methodological and requires no additional data. Grouped ablation, in which all pressure channels or all temperature channels are withheld together, would separate the value of a physical quantity from the value of one sensor reporting it, which single-feature ablation cannot do. A forward-selection counterpart, adding signals one at a time to the static model, would show how quickly the achievable performance is reached. Calibrating the ensemble output itself, rather than only its component inputs, would address the calibration error reported in Section 4. Reporting attributions computed on the ensemble, through permutation importance or SHAP values, together with the stability of each rank across folds, would align the model that is explained with the model that is evaluated. Reporting AUPRC alongside AUC would describe performance more faithfully at an event rate of 11.6%.

A second group of directions requires more data. External validation on larger, multi-center cohorts would test generalizability, and the finding that registry variables alone suffice makes such validation markedly easier to organize. With a larger case count, the composite endpoint could be decomposed, so that prolonged ventilation, renal failure and reoperation for bleeding are predicted separately, which would yield risk estimates that map to specific preventive actions rather than to a single undifferentiated alert. Architectures designed for multivariate clinical time series could then be evaluated on their merits rather than being constrained by sample size, and pretraining a sequence encoder on unlabeled operations, which are abundant where labeled adverse events are not, is a plausible route to making the deep branch contribute.

A third group concerns the data elements not yet included. Real-time intraoperative medication data, including continuous infusion rates of vasopressors, inotropes, anesthetic medications, intravenous fluid, blood products and coagulation factors, would capture pharmacologic management, which is among the most directly modifiable aspects of intraoperative care and therefore among the most likely to add information that the registry does not already encode. Serial intraoperative laboratory measurements such as arterial blood gases, lactate trends and coagulation parameters would track metabolic derangement as it develops. Beyond the eleven STS-defined events examined here, postoperative delirium, which affects up to 50% of cardiac surgery patients and is associated with increased mortality [42], is an important additional target, as are graft patency and longer prediction horizons at 90 days, one year and five years.

A fourth group concerns deployment. A model that is never seen by a clinician changes nothing, and the work on usability and implementation of comparable screening tools shows that adoption depends on how output is presented and on whether it fits existing workflow [11, 12, 38]. Developing an interface in which the care team can see a risk score at handoff, together with the factors driving it and the response to treatment over time, is therefore not a secondary engineering task but a condition of clinical impact.

### 7.3 Vision

The broader aim we are working toward is that risk assessment in cardiac surgery should be continuous rather than a single number computed before the incision and never revised. A patient’s risk changes during the operation and again during the first hours in the ICU, and the information that would support revising it is already being recorded. The handoff from operating room to intensive care unit is the point in that trajectory where information is most reliably lost [40], and it is the point this study targets.

Our results suggest a more tempered version of that vision than we began with. On this cohort the intraoperative record did not add discriminative power beyond the registry, which indicates that the value of continuous assessment will not come from waveform data alone. It is more likely to come from the elements not yet in the model, particularly pharmacologic management and serial laboratory values, which reflect what the care team actually does rather than restating how sick the patient already was. A realistic path is therefore incremental: establish that a registry-based model generalizes across institutions, add the intraoperative elements that carry genuinely new information, decompose the endpoint as the case count allows, and put the result in front of clinicians in a form they can act on.

If that path is followed, the contribution of models such as this one will be measured not by AUC but by whether complications are recognized earlier than they otherwise would be, and by whether the variation in failure to rescue between institutions [9, 23] narrows. That is the standard against which we would wish this line of work to be judged.

## Declaration on the Origin of Figures

All figures in this manuscript were produced by the authors. Figure 1 is a schematic of the model architecture drawn by the authors in Google Slides. Figures 2 to 10 were generated in Python directly from the study data by the authors’ analysis code, and each plots values computed in the cross-validation described in Section 2. No figure in this manuscript was generated, synthesized or stylistically altered by a generative artificial intelligence system, and no figure contains simulated or illustrative data.

## Declaration on the Use of Artificial Intelligence in Writing

The authors used a generative artificial intelligence assistant during the preparation of this manuscript for language editing purposes. The authors take full responsibility for the content, accuracy and integrity of the manuscript.

## Author Contributions

Conceptualization, S.A. and A.H.; methodology, S.A. and R.K.; software, S.A. and K.R.; validation, S.A., K.R., A.H.; formal analysis, S.A. and K.R.; investigation, S.A., R.K. and D.K.; resources, S.A., K.R., A.H., R.K., D.S., R.W., F.M. and D.K.; data curation, K.R.; writing, original draft preparation, S.A. and K.R.; writing, review and editing, S.A., R.K., A.H., F.M., T.K., R.K., Q.J., D.S., R.W., M.M.; visualization, R.K.; supervision, S.A.; project administration, S.A. All authors have read and agreed to the published version of the manuscript.

## Data Availability

The data used in this study were obtained from the Society of Thoracic Surgeons Adult Cardiac Surgery Database (STS ACSD) records and intraoperative monitoring data of patients treated at MaineHealth Maine Medical Center. These data contain protected health information and are subject to institutional data use agreements and STS participation terms, so they cannot be shared publicly. De-identified data may be made available from the corresponding author on reasonable request, subject to approval by MaineHealth and execution of a data use agreement. The analysis code is available from the corresponding author on reasonable request.

